# 58 risk and 34 protective effects of *GLP1R* on disease phenome and adverse neonatal health

**DOI:** 10.64898/2026.01.24.26344404

**Authors:** Robert H. Campbell, Melinda C. Mills

## Abstract

Cardiometabolic gene-target drugs such as Glucagon-Like Peptide 1 Receptor agonists are used extensively, yet risks and repurposing have not been systematically evaluated across a comprehensive set of diseases. We use Phenome-wide Mendelian Randomization to test *GLP1R* gene expression and Cholesteryl Ester Transfer Protein (CETP) concentration, two promising cardiometabolic drug targets, on risk of 396 diseases based on sex-specific, ancestry-specific, genome-wide association studies in UK Biobank. We identify 92 causal effects (66 novel) of genetically proxied *GLP1R* expression on disease, with 58 protective and 34 risk effects. Risks of *GLP1R* expression on neonatal health raise concern for women who may become pregnant. *GLP1R* expression substantially increases risk of Vitamin D deficiency.

Although *GLP1R* is hypothesized as anti-ageing, we find it increases risk of 22 age-related diseases. Conversely, we find CETP inhibition is narrowly cardioprotective. Results show benefits, risks and repurposing opportunities for *GLP1R*-targetted and CETP-targeted drugs.

## Introduction

Cardiovascular diseases are the leading cause of death worldwide^1^, while prevalence of metabolic diseases such as type 2 diabetes has more than doubled since 1990, accounting for 10% of the annual UK National Health Service budget^2^. Multiple gene-target drugs have been developed to counter cardiometabolic disease, creating optimism and concern about rapid adoption. The most prominent are GLP-1R agonists, a class of drugs originally approved to treat diabetes. GLP-1R drugs target the GLP-1 receptor, which is encoded by the eponymous *GLP1R* gene. As of January 2026, GLP-1R agonist drugs are used by as many as one in eight Americans to treat overweight or obesity^3^. Due to upcoming patent expiry, the ongoing development of oral formulations, and plans to lower prices, GLP-1R agonist use is expected to further increase^4^, meriting both optimism and caution. Recent observational evidence suggests that GLP-1R agonists may have additional risks and benefits outside their original target indications^5^, including possibly as anti-ageing drugs^6^.

Like the former morning-sickness drug thalidomide^7^, GLP-1R agonists are untested for pregnancy safety but increasingly used on reproductive-aged women. However, GLP-1 has been shown to cause birth complications and fetal death in mouse models^8,9^. This presents a public health concern for women who may become pregnant given the multi-week half-life of some GLP-1R agonists^10^, and the fact that approximately 50% of pregnancies are unplanned^11^.

Cholesteryl ester transfer protein (CETP) inhibitors are a second class of gene-target cardiometabolic drugs promising to disrupt the current best-in-class lipid lowering therapy (PCSK-9 inhibitors) with an oral formulation. CETP inhibitors are hypothesized to reduce cardiovascular disease by modifying multiple blood lipid parameters (LDL-C, Apo-B, HDL-C) including LP(a), a highly atherogenic particle that until recently was believed to be unmodifiable through lifestyle or pharmacology^12^.

The CETP inhibitor drug Obicetrapib is currently under European Medicines Agency (EMA) review for treatment of high cholesterol^13^. However, previous CETP inhibitor drug programs, such as Torcetrapib, have been terminated due to the discovery of elevated blood pressure and increased deaths in the treatment arm, raising concerns about side effects^14^. It is also unknown whether CETP inhibition could modify risk of diseases beyond high cholesterol.

Due to the genetic mechanism of action of these drug classes, causal genomic analysis offers a unique opportunity to address the limitations in existing literature from clinical trial results, animal data, and observational research. Drug-targeting Phenome-wide Mendelian Randomization (MR-PheWAS) is a causal statistical technique that performs a “phenome scan” for the effect of perturbations of a genetic drug target on the phenome of human disease^15^.

Unlike a traditional phenome scan, the drug target exposure is estimated using meta-analysis of quantitative trait loci (QTLs) of the drug target gene, and exposure-outcome relationships are estimated using the Mendelian Randomization method, which leverages the random-like assignment of alleles to estimate causal effects^16,17^.

MR-PheWAS allows for natural-experiment tests of gene activity levels using high sample sizes in a human population over the course of an entire lifetime^17,18^, and can test hypotheses in vulnerable populations such as pregnant women, where assignment to a treatment group would be unethical.

As of January 19^th^ 2026, a PubMed search shows no phenome-wide causal research of either *GLP1R* or CETP (Supplementary Methods, Prior literature search). We found 44 and 75 individual-phenotype Mendelian randomization studies of *GLP1R* and CETP respectively, which test for the effect of these drug targets on individual “candidate diseases”^19^ rather than the full phenome of disease. These findings have mostly been in the protective direction. Genetic analysis of repurposing potential using MR-PheWAS can validate and expand on existing research^5^, and test for unexamined risks and repurposing opportunities, such as for neonatal diseases.

We used MR-PheWAS to investigate the causal effects of genetically proxied CETP concentration and *GLP1R* expression on a broad range of diseases, with a particular focus on neonatal diseases, age-related diseases, and sex differences.

Genetic instruments for CETP concentration and *GLP1R* expression were derived from published genome-wide, protein-quantitative, and expression-quantitative trait loci studies, using sex-specific instruments where available (CETP) and both-sex instruments where genetic effects were assumed to be sex-conserved^20^ (*GLP1R*). Outcomes were drawn from hundreds of disease phenotypes defined using Global Burden of Disease (GBD)^21^ hierarchies and mapped to ICD-10 codes with genome-wide association study (GWAS) summary statistics from two pan-phenome GWAS of the UK Biobank^22,23^, analyzed separately in males, females, and both sexes. The combination of these powerful data sources allows for methodologically rigorous coverage of the human disease phenome, combined with the high statistical power of population-scale genomic data applied to granular, ICD-10-specific phenotyping.

Our MR-PheWAS used standard instrument quality control^16^ and MR estimators (Wald ratio for single-instrument exposures, inverse-variance weighted methods for multi-instrument exposures)^24^, followed by extensive robustness checks to mitigate bias from pleiotropy, heterogeneity, and reverse causation^16^. Significant exposure–outcome associations were identified after Benjamini-Hochberg false discovery rate correction (q < 0.05). We included positive and negative outcome controls for both *GLP1R* (type 2 diabetes (E11) and hair color^25^) and CETP (hypercholesterolemia^26^ and hair color^25^) to validate our instruments, as well as positive and negative exposure controls (smoking ^27–29^ and handedness^22^) to test our MR-PheWAS pipeline.

To identify the risk or protective effects of *GLP1R* and CETP on disease categories, we grouped our MR-PheWAS results using the GBD disease ontology and performed permutation tests on in-group versus out-group disease signed p-values. We also applied permutation testing to compare MR effects across age-related diseases (ARDs) versus non-age-related diseases^30^.

Sensitivity analyses examined sex-conserved *GLP1R* effects, sex-divergent *GLP1R* effects, and maternal and neonatal disease subsets. The study was conducted in accordance with STROBE-MR and established MR guidelines, adapting them for a hypothesis-free, phenome-wide context^16,31^.

In this work, we show *GLP1R* expression protects against 58 ICD-10-coded diseases, while raising risk for 34 others. Of particular relevance are the risk effects of *GLP1R* expression on ICD-10 coded neonatal health outcomes. We find CETP inhibition has an expected protective effect on cardiovascular disease, with no other significant effects.

## Results

### *GLP1R* – MR-PheWAS

Our MR-PheWAS analyses (Figure 1) of *GLP1R* expression show phenome-wide significant effects across all categories of diseases, both protective and with risks, for women, men, and both sexes. While we observe differences in the proportions of risk and protective effects by disease category, permutation testing for category-wide effects was null after multiple testing correction.

**Figure 1.**
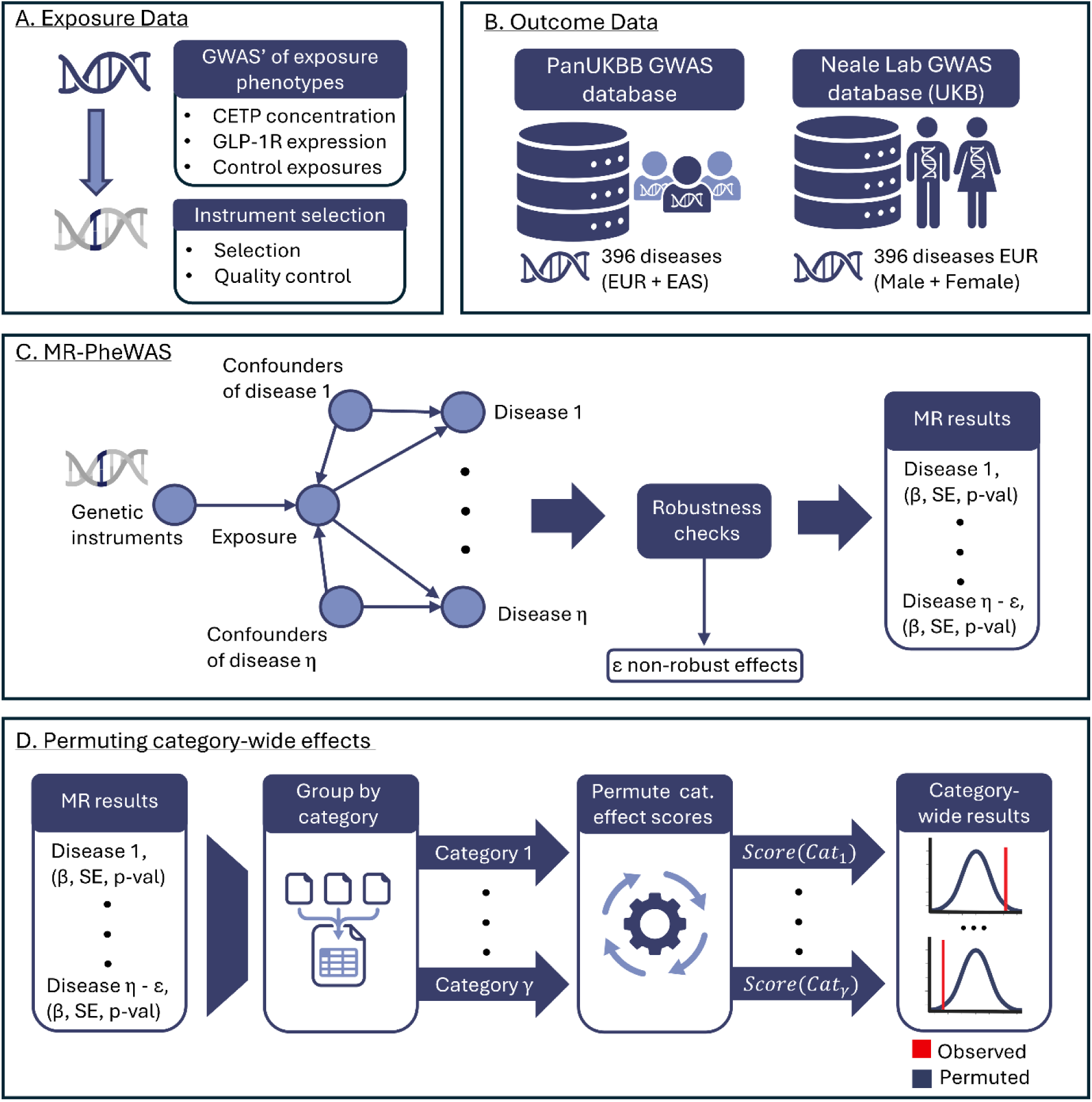
Overview of model design. Study design and analysis workflow for MR-PheWAS. **(A)** Exposure data comprised quantitative trait loci of CETP concentration and *GLP1R* gene expression filtered for suitability in MR-PheWAS using standard thresholds. **(B)** Outcome data comprised the Genome-Wide Association Study summary statistics for 396 traits at the selected genomic loci from (A) made available through the PanUKBB and Neale Lab UKB GWAS Round 2 projects. **(C)** MR-PheWAS main model. The causal effect of the exposure from (A) is estimated on each outcome selected from (B) using two-sample MR. Robustness checks and multiple testing correction filter out non-robust results. **(D)** The estimated causal effects from (C) are grouped by disease category and permutation tested for category-wide effects of the exposure.

For the MR-PheWAS of *GLP1R*, we used a shared set of single nucleotide polymorphisms (SNPs) available through eQTLGen as exposure instruments for *GLP1R* gene expression^32,33^ in females, males and both sexes (Table 1, Supplementary Table 1). 22 of 134 candidate *GLP1R* SNPs passed exposure quality control and were used as exposure instruments for MR-PheWAS (Supplementary Table 2).

**Table 1.** MR-PheWAS summary results for CETP, *GLP1R* and control exposures on sex-specific, ancestry-specific populations.

| Exposure | Ancestry | Sex | Pleiotropy Screening Regex | Exposure Units | Instr. p-value threshold | Instr. count | Diseases passing robustness check (ARD/non-ARD) | Significant disease effects (risk/protection) | Category-wide effects |
| --- | --- | --- | --- | --- | --- | --- | --- | --- | --- |
| *GLP1R* | EUR | Both | None | SD *GLP1R* gene expression | 5x10<sup>-8</sup> | 22 | 148 (89/59) | 92 (34/58) | FALSE |
|  | EUR | Female | None | SD *GLP1R* gene expression | 5x10<sup>-8</sup> | 22 | 345 (170/175) | 213 (101/112) | FALSE |
|  | EUR | Male | None | SD *GLP1R* gene expression | 5x10<sup>-8</sup> | 22 | 298 (207/92) | 202 (88/114) | FALSE |
| CETP | EUR | Both | None | SD CETP protein concentration | 5x10<sup>-8</sup> | 3 | 347 (197/150) | 0 | Cardiovascular disease, level two |
|  | EUR | Female | None | SD CETP gene expression | 5x10<sup>-2</sup> ** | 1 | 345 (170/175) | 0 | NA |
|  | EUR | Male | None | SD CETP gene expression | 5x10<sup>-4</sup>* | 1 | 304 (210/94) | 0 | NA |
| Left-handed | EUR | Both | None | log-OR left handed | 5x10<sup>-8</sup> | 2 | 0 | NA | NA |
| Ambidextrous | EUR | Both | None | log-OR ambidextrous | 5x10<sup>-8</sup> | 1 | 118 (68/50) | 0 | NA |
| Smoking initiation | EUR | Both | none | log-OR ever smoker | 5x10<sup>-8</sup> | 92 | 300 (180/120) | 50 (50/0) | FALSE |
|  | EUR | Both | "educat", "school", "BMI", "Obes" | log-OR ever smoker | 5x10<sup>-8</sup> | 1 | 152 (91/61) | 0 | NA |
|  | EAS | Both | None | log-OR ever smoker | 5x10<sup>-8</sup> | 4 | 20 (12/8) | 0 | FALSE |
|  | EAS | Male | "educat", "school", "BMI", "Obes" | log-OR ever smoker | 5x10<sup>-8</sup> | 0 | NA | NA | NA |
|  | EAS | Female | "educat", "school", "BMI", "Obes" | log-OR ever smoker | 5x10<sup>-8</sup> | 0 | NA | NA | NA |

\* Benjamini-Hochberg false discovery rate adjusted significant at $q < 0.05$
\*\* F statistic is lower than 10.

Of the 321 diseases in the both-sex phenome with available GWAS data included in the both-sex MR-PheWAS of *GLP1R*, 148 (89 ARD and 59 non-ARD) passed our robustness checks (Figure 2, Supplementary Table 3). Of the 148 remaining results shown in Figure 2, *GLP1R* had a phenome-wide significant effect on 92 diseases defined by ICD-10 codes (q < 0.05), with a protective effect on 58 and a risk effect on 34.

**Figure 2.**
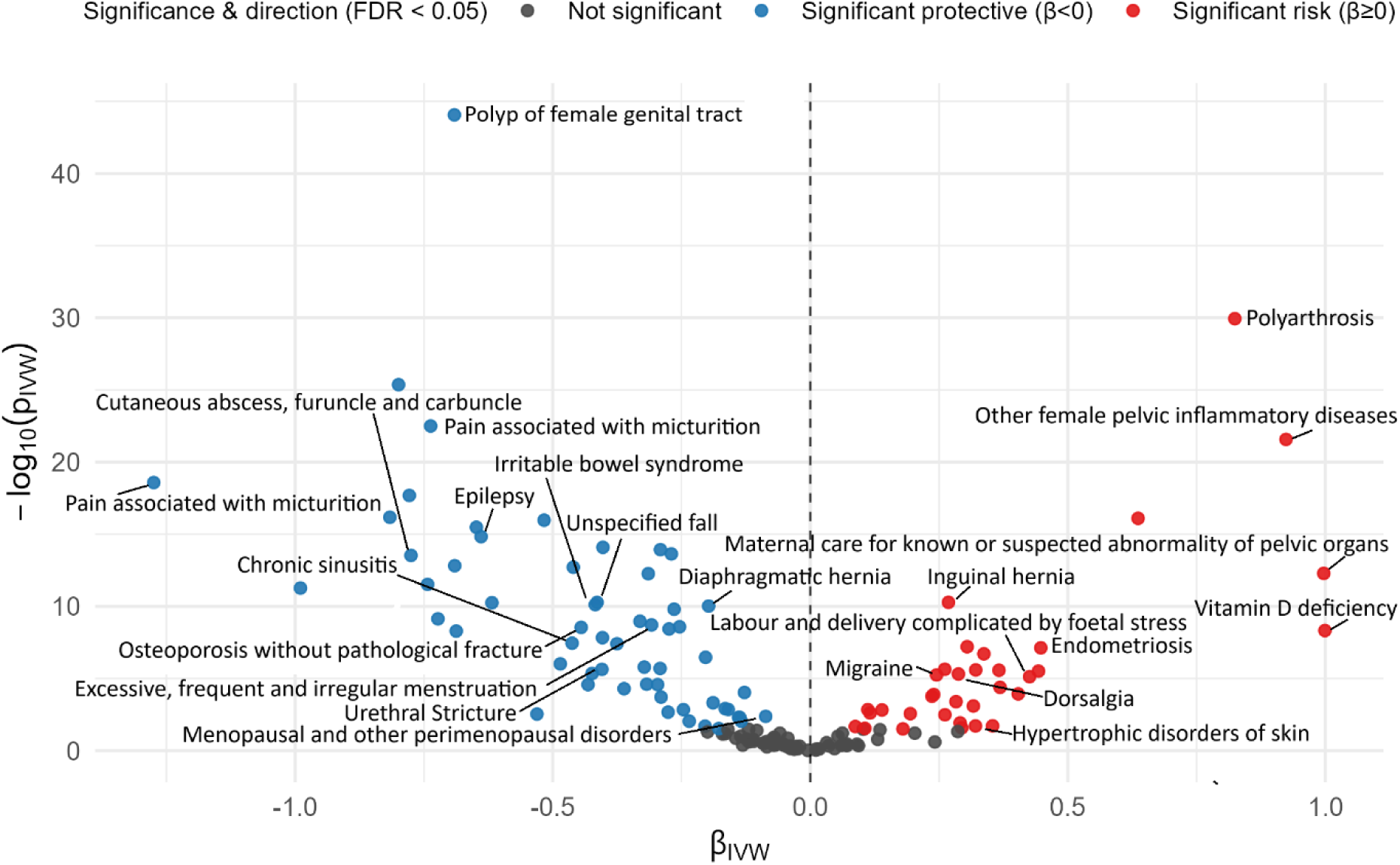
Volcano plot results of phenome-wide Mendelian randomization of *GLP1R* expression. “MR” refers to Mendelian Randomization testing. “*GLP1R*” refers to glucagon-like peptide 1 receptor gene expression. “FDR” refers to Benjamini-Hochberg false discovery rate correction. “βivw” refers to the causal effect of one standard deviation increase of *GLP1R* expression on log-odds ratio of disease diagnosis as estimated using the inverse variance weighted method. The dotted line refers to a log-odds ratio of zero, implying no effect change in disease risk resulting from increased *GLP1R* expression. Disease labels are applied to selected novel Mendelian randomization findings with selective labelling. Red dots imply increased *GLP1R* expression raises risk for the named disease outcome, while blue dots imply increased *GLP1R* expression lowers risk. Statistical significance is defined as q < 0.05 after Benjamini-Hochberg false discovery rate adjustment.

The disease with the largest protective effect size in the both-sex *GLP1R* MR-PheWAS was “Pain associated with micturition” (OR 0.28), followed by Cardiac arrest (OR 0.37). The 10 largest protective effect size diseases included common healthspan-ending diseases, such as cardiovascular diseases (I46, I49), cancers (C79) and neurodegenerative conditions (G81).

The disease with the largest risk effect size in the both-sex *GLP1R* MR-PheWAS was Vitamin D deficiency, with an odds ratio of 2.72 per standard deviation increase in *GLP1R* expression, followed by “Maternal care for known or suspected abnormality of pelvic organs” (OR ≈ 2.72), “Other female pelvic inflammatory diseases” (OR 2.51), and “Polyarthrosis” (OR 2.28). Four of the 10 diseases with the highest risk were related to the uterus or female reproduction; others included “Endometriosis” (OR 1.55) and “Labor and delivery complicated by fetal stress” (OR 1.55). We note that the naïve odds ratios for female-specific diseases are likely to underestimate the true risk effect, as they are estimated over a sample population containing approximately 46% men, who are not in the risk set. We also note that these effect sizes reflect the effect of natural variation of *GLP1R* expression levels among people, not the considerably larger effect of GLP-1R agonist use (see Discussion).

We conducted literature searches for existing MR studies of *GLP1R* to identify whether our 92 significant results for the both-sex population contained novel findings. We performed a primary search using PubMed for each of our significant results, checking against Perplexity^34^ if the PubMed search returned no relevant results (Supplementary Methods, Literature Search for Novel Findings). We found no prior MR studies for 66 of our 92 significant findings. Of these 66 novel MR findings, 41 were protective findings, where *GLP1R* reduced risk of an ICD-10-coded disease, and 25 were risk findings, where *GLP1R* increased risk of an ICD-10-coded disease (Supplementary Table 4).

Results from the MR-PheWAS of *GLP1R* in the both-sex population grouped by Global Burden of Disease level-2 categories are shown in Figure 3. The most protective-skewed category was the expected “Diabetes and kidney diseases,” and the most risk-skewed category was “Musculoskeletal disorders.” Our sensitivity check of “sex-conserved” findings that treated the female population as discovery and male population as validation returned a set of 98 phenome-wide significant effects of *GLP1R* (Supplementary Table 5). Our sensitivity check of “sex-divergent” findings returned a set of 56 statistically significant sign disagreements between the effects of *GLP1R* on males and females (Supplementary Table 6).

**Figure 3.**
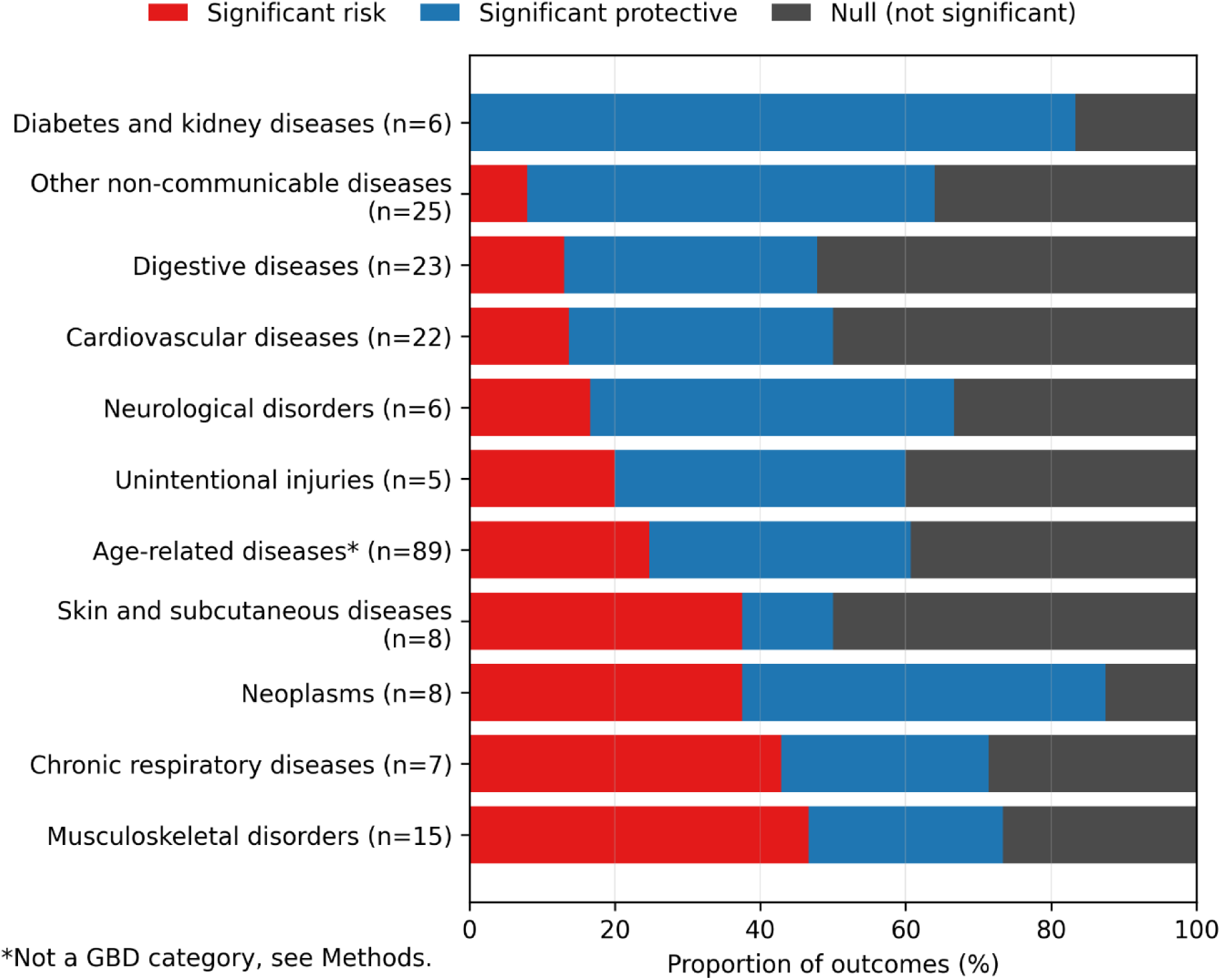
G*L*P1R disease effects grouped by Global Burden of Disease level-2 category (both-sex population). Colored bars represent categories of disease drawn from level two of the Global Burden of Disease ontology. The bracketed number (n = X) next to the category name represents the number of ICD-10 coded diseases tested in the MR-PheWAS of *GLP1R* expression mapped to that category. The red colored portion of each bar represents the proportion of diseases sorted into that category on which increased *GLP1R* expression has robust, phenome-wide significant risk effect. The blue colored portion of each bar represents the proportion of diseases sorted into that category on which increased *GLP1R* expression has a robust, phenotype-wide significant protective effect. The gray colored portion of bar represents the proportion of diseases sorted into that category on which increased *GLP1R* expression does not have a directionally significant effect. Categories marked with an asterisk were not derived from the Global Burden of Disease ontology. Bars are sorted from the lowest proportion of (red) risk effects to the highest. Categories with fewer than five mapped diseases are excluded.

### *GLP1R* – MR-PheWAS – Sex-specific

Of the 345 ICD-10 codes in the female phenome with available GWAS surviving robustness checks (170 ARD, 175 non-ARD), *GLP1R* expression had a statistically significant effect on 213 (101 risk; 112 protective) after false discovery rate correction (q < 0.05) (Supplementary Table 7, Supplementary Figure 1). Of the 298 ICD-10 codes with available GWAS in the male phenome surviving robustness checks (207 ARD, 92 non-ARD), *GLP1R* expression had a statistically significant effect on 202 diseases (88 risk; 114 protective) (Supplementary Table 8, Supplementary Figure 2). These results show significant effects of *GLP1R* on 64% and 67% of the phenotypes tested respectively.

To investigate the effects of *GLP1R* on pregnancy, we examined the proportions of risky, protective and null effects of *GLP1R* on the categories of “neonatal diseases,” which describe pregnancy risks originating from the neonate, and “maternal diseases,” which describe pregnancy risks originating from the mother (see Methods, Maternal and neonatal diseases, and Supplementary Table 9). Table 2 shows the estimated effects of *GLP1R* on individual disease effects in both categories as measured in the female population. We found “neonatal diseases” was the most risk-affected category of the 14 level-2 disease categories examined (Figure 4).

**Figure 4.**
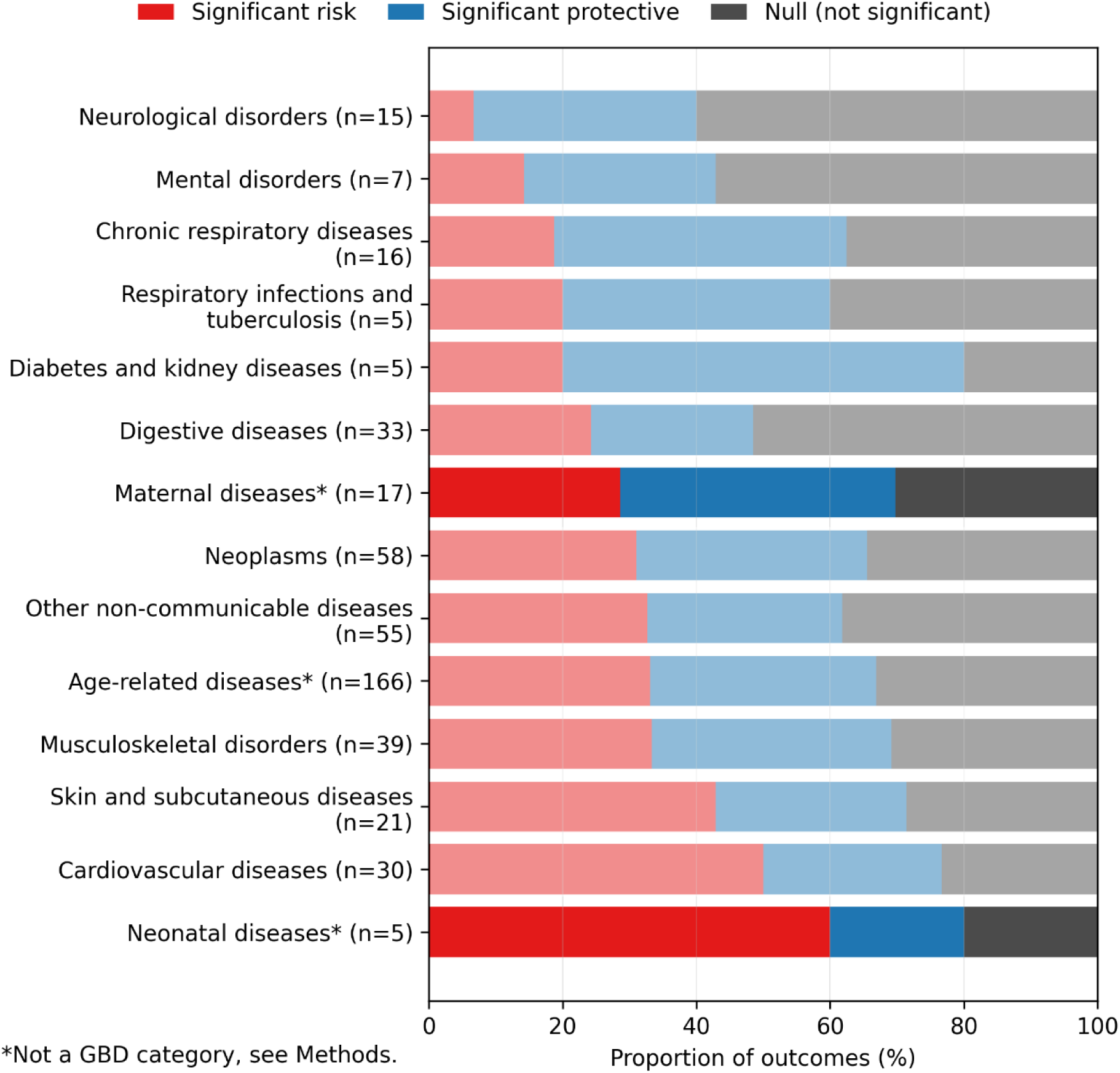
female sex-specific *GLP1R* disease effects grouped by Global Burden of Disease level-2 category (maternal and neonatal diseases highlighted). Colored bars represent categories of disease, with maternal and neonatal diseases highlighted. The bracketed number (n = X) next to the category name represents the number of ICD-10 coded diseases tested in the MR-PheWAS of *GLP1R* expression mapped to that category. The red colored portion of each bar represents the proportion of diseases sorted into that category on which increased *GLP1R* expression has robust, phenome-wide significant risk effect. The blue colored portion of each bar represents the proportion of diseases sorted into that category on which increased *GLP1R* expression has a robust, phenotype-wide significant protective effect. The gray colored portion of bar represents the proportion of diseases sorted into that category on which increased *GLP1R* expression does not have a directionally significant effect. Categories marked with an asterisk were not derived from the Global Burden of Disease ontology. Bars are sorted from the lowest proportion of (red) risk effects to the highest. Categories with fewer than five mapped diseases are excluded.

**Table 2.** Estimated directional risk and protective effects of increased *GLP1R* expression on maternal and neonatal diseases in female European population.

| Maternal Diseases | Naïve p-value | BH-adjusted q-value | Robustness checks (passed / attempted) | Phenome-wide significant | Classification |
| --- | --- | --- | --- | --- | --- |
| O14 Gestational [pregnancy-induced] hypertension with significant proteinuria | 3.67E-06 | 1.19E-05 | (7/7) | TRUE | Significant risk |
| O16 Unspecified maternal hypertension | 0.015 | 0.03 | (7/7) | TRUE | Significant risk |
| O69 Labor and delivery complicated by umbilical cord complications | 0.025 | 0.04 | (7/7) | TRUE | Significant risk |
| O72 Postpartum hemorrhage | 2.02E-05 | 5.78E-05 | (7/7) | TRUE | Significant risk |
| O20 Hemorrhage in early pregnancy | 2.98E-06 | 9.88E-06 | (7/7) | TRUE | Significant protective |
| O21 Excessive vomiting in pregnancy | 7.38E-04 | 1.50E-03 | (7/7) | TRUE | Significant protective |
| O48 Prolonged pregnancy | 2.87E-03 | 5.40E-03 | (7/7) | TRUE | Significant protective |
| O75 Other complications of labor and delivery, not elsewhere classified | 1.44E-20 | 4.03E-19 | (7/7) | TRUE | Significant protective |
| O42 Premature rupture of membranes | 8.05E-06 | 2.41E-05 | (7/7) | TRUE | Significant protective |
| O46 Antepartum hemorrhage, not elsewhere classified | 8.68E-05 | 2.15E-04 | (7/7) | TRUE | Significant protective |
| O64 Obstructed labor due to malposition and malpresentation of fetus | 0.031 | 0.05 | (7/7) | TRUE | Significant protective |
| O03 Spontaneous abortion | 0.046 | 0.07 | (7/7) | FALSE | Not significant |
| O13 Gestational [pregnancy-induced] hypertension without significant proteinuria | 0.634 | 0.68 | (7/7) | FALSE | Not significant |
| O44 Placenta previa | 0.265 | 0.33 | (7/7) | FALSE | Not significant |
| O62 Abnormalities of forces of labor | 0.433 | 0.49 | (7/7) | FALSE | Not significant |
| O63 Long labor | 0.118 | 0.16 | (7/7) | FALSE | Not significant |
| O70 Perineal laceration during delivery | 0.145 | 0.19 | (7/7) | FALSE | Not significant |

| Neonatal Diseases | Naïve p-value | BH-adjusted q-value | Robustness checks (passed / attempted) | Phenome-wide significant | Classification |
| --- | --- | --- | --- | --- | --- |
| O68 Labor and delivery complicated by fetal stress [distress] | 0.022 | 0.04 | (7/7) | TRUE | Significant risk |
| O69 Labor and delivery complicated by umbilical cord complications | 0.025 | 0.04 | (7/7) | TRUE | Significant risk |
| O00 Ectopic pregnancy | 7.07E-09 | 3.29E-08 | (7/7) | TRUE | Significant risk |
| O02 Other abnormal products of conception | 2.35E-09 | 1.14E-08 | (7/7) | TRUE | Significant protective |
| O03 Spontaneous abortion | 0.046 | 0.07 | (7/7) | FALSE | Not significant |

For neonatal diseases, *GLP1R* had a 25% protective and 75% risky effect among the four phenome-wide, significantly affected diseases within-category. The three significant risk effects on neonatal diseases were “O69 Labor and delivery complicated by umbilical cord complications,” “O00 Ectopic pregnancy,” and “O68 Labor and delivery complicated by fetal stress” (Table 2). O86 previously appeared as a risk effect in the both-sex MR-PheWAS of *GLP1R*. The single significant protective effect on the neonatal phenome was “O02 Other abnormal products of conception”. Conversely, *GLP1R* had a 63% protective and 37% risky effect on the 11/17 significantly affected maternal diseases.

### *GLP1R* Age-Related Disease

*GLP1R* had at least one significant risk and protective effect in every level-2 GBD disease category containing age-related diseases in the both-sex, female, and male MR-PheWAS (Supplementary Figures 3-5) and corresponding sex-specific ageing phenomes (Supplementary Tables 10-12). Figure 5 shows phenome-wide significant risk (22) and protective (32) effects for 54 of the 89 diseases (60%) in the both-sex age-related disease phenome. *GLP1R* showed a concentrated protective effect on diabetic and kidney diseases within the ageing phenome, where it significantly protected against 5 of 6 diseases in this category (Figure 5). Conversely, *GLP1R* showed a mixed-to-risky effect on musculoskeletal disorders in the ageing phenome, where it had a significant risk effect on 6 diseases and a protective effect on 4. Permutation tests for an ageing-phenome-wide effect of *GLP1R* were null for women, men and both sexes (Supplementary Figures 6-8).

**Figure 5.**
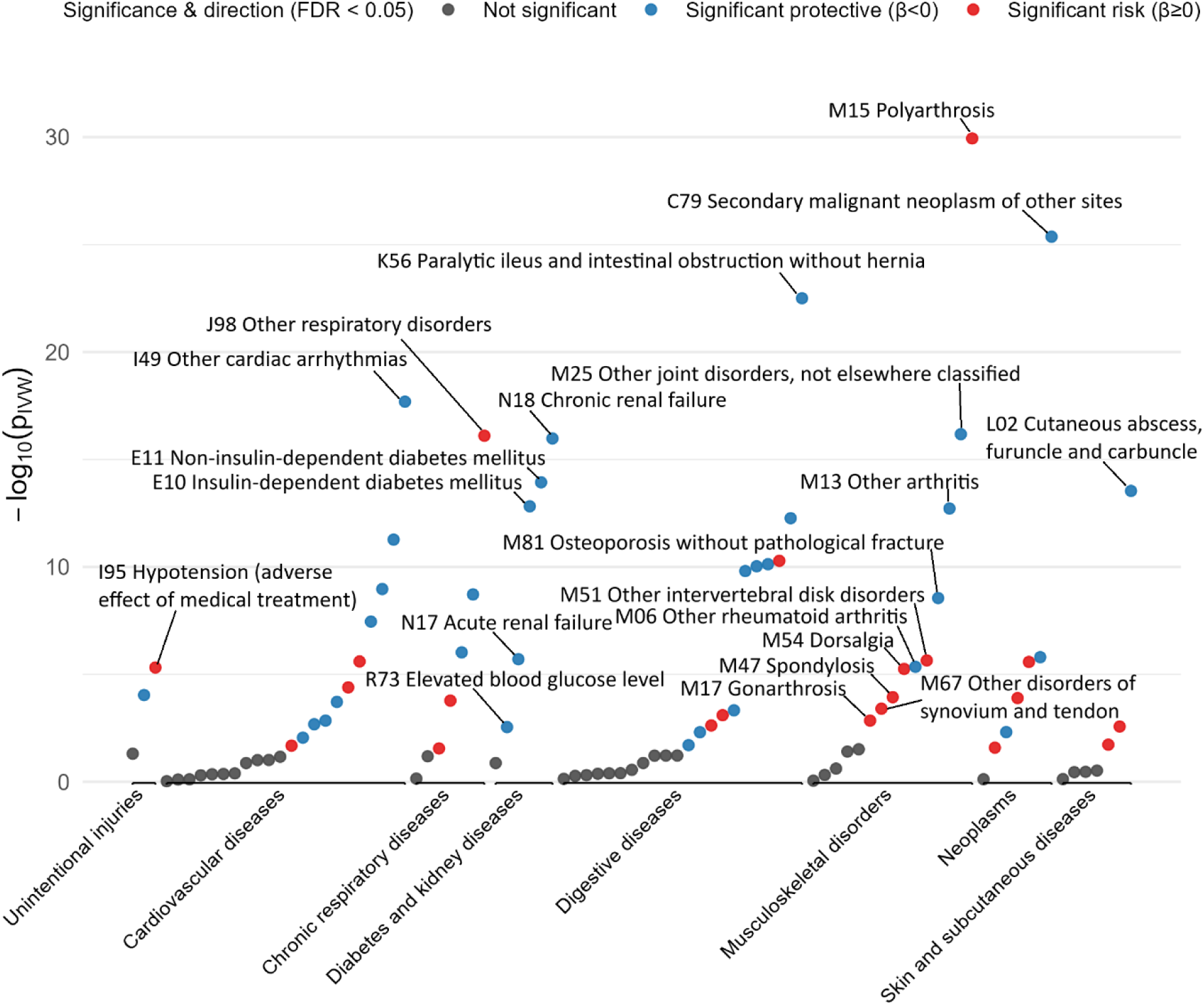
Manhattan plot of *GLP1R* effects on the phenome of age-related diseases in the both-sex population. Diseases are grouped by Global Burden of Disease category (level two). Disease labels are applied to the lowest p-value effect in each category, and to significantly affected diseases in the most risk-affected and protective-affected categories. Red dots imply increased *GLP1R* expression raises risk for the named disease outcome, while blue dots imply increased *GLP1R* expression lowers risk. Statistical significance is defined as q < 0.05 after Benjamini Hochberg false discovery rate adjustment.-log10(Pivw) represents the log-scaled p-value measuring the statistical significance of the measured effect.

### *GLP1R* – Disease Category Effect

To test for the statistical significance of the observed disease-group differences, we conducted permutation testing for a category-wide risk or protective effect of *GLP1R* on all disease categories from the full GBD disease ontology using the top three disease hierarchy levels^21^, including the three additional categories “neonatal diseases,” “maternal diseases,” and “age-related diseases.” After accounting for multiple testing burden, we found no category-wide significant results at any level for both-sex (Supplementary Figures 9-11), female (Supplementary Figures 12-14) and male (Supplementary Figures 15-17) analyses (q < 0.05).

These permutation tests were adjusted for multiple testing burden (q < 0.05) at the disease hierarchy level (1,2,3). Relaxing multiple testing correction for the both-sex analyses results in significant group-wide protection of *GLP1R* on “neoplasms” (level two, p = 0.031), and “other neoplasms” (level three, p = 0.026).

### CETP – MR-PheWAS

The results for the MR-PheWAS of CETP protein concentration show a significant protective effect of CETP inhibition on cardiovascular disease at both the individual disease and disease-group level, with no other significant effects (Table 1, Supplementary Tables 1,15).

We used exposure SNPs from a GWAS of CETP concentration in our both-sex analysis, and a single sex-specific CETP concentration SNP for the sex-specific analyses^35,36^ (Supplementary Tables 13-14). For the both-sex analysis shown in Figure 6, the MR-PheWAS pipeline fit IVW results for the 396 diseases with available GWAS data (224 ARD; 172 non-ARD) in UK Biobank, of which 347 passed robustness checks (197 ARD; 150 non-ARD) (Supplementary Table 15). Of these, we identified a phenome-wide significant risk effect of increased CETP concentration on Angina Pectoris (I20), an acute ischemia of the heart resulting in chest pain, typically caused by coronary artery disease.

**Figure 6.**
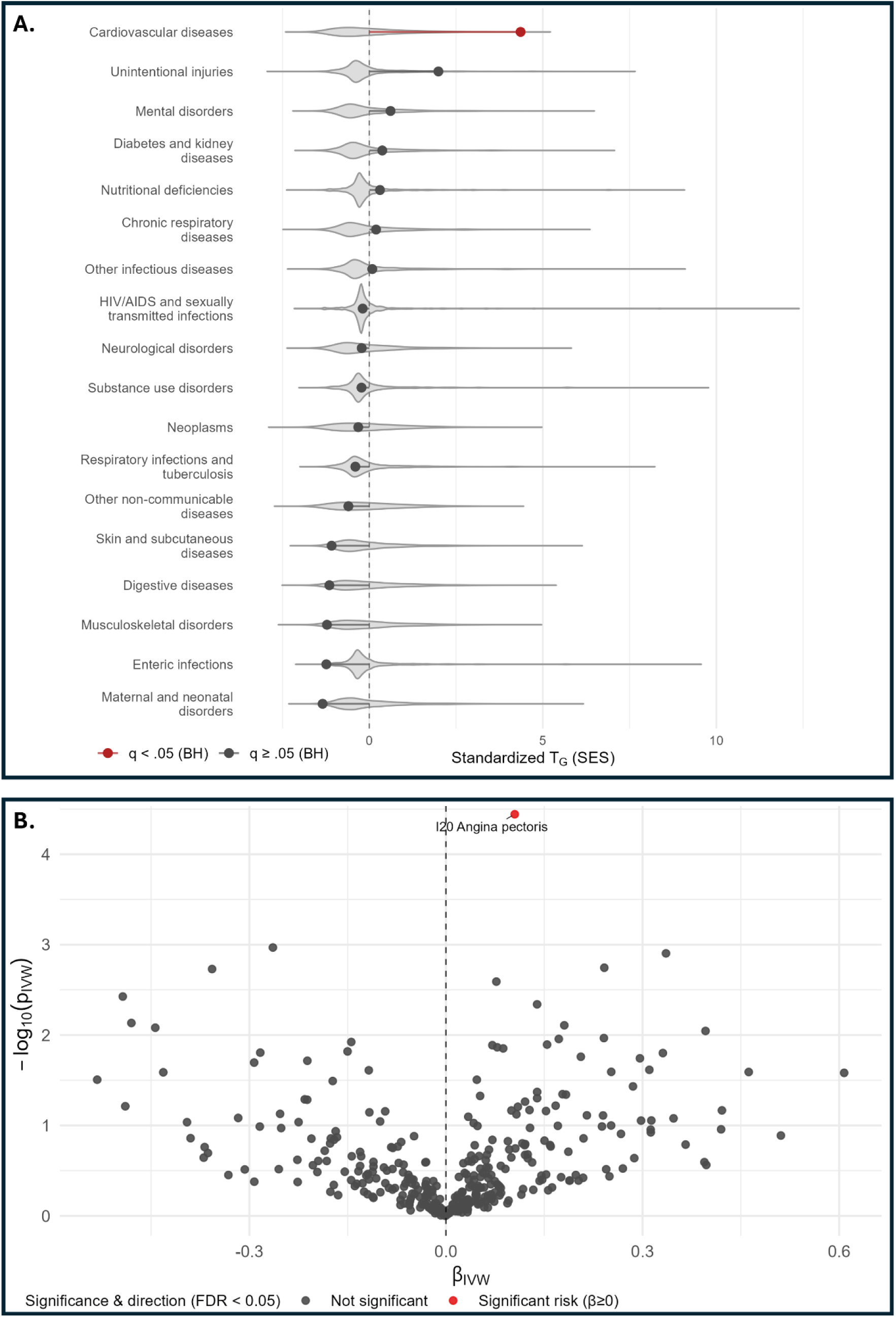
Summary effects of increased CETP concentration. **(A)** Permutation testing for category-wide effects of CETP concentration on the European both-sex population. Categories are drawn from level two of the Global Burden of Disease clinical ontology. Red dots imply increased CETP concentration raises risk for the diseases in the named disease category. Statistical significance is defined as q < 0.05 after Benjamini-Hochberg false discovery rate adjustment over the 18 categories at level two. **(B)** Volcano plot results of phenome-wide Mendelian randomization of CETP concentration in the both-sex European population. “MR” refers to Mendelian Randomization testing. “CETP” refers to cholesteryl ester transfer protein. “FDR” refers to Benjamini-Hochberg false discovery rate correction. “βivw” refers to the causal effect of one standard deviation increase of CETP concentration in whole blood on the log-odds ratio of disease diagnosis as estimated using the inverse variance weighted method. The dotted line refers to a log-odds ratio of zero, implying no effect change in disease risk resulting from increased CETP concentration.

Because the sex-specific MR-PheWAS of CETP used only a single exposure instrument, we used the Wald ratio as our primary MR estimator and disabled the robustness analysis pipeline, which is undefined on single-instrument analyses. For the female analysis, the MR-PheWAS pipeline fit Wald ratio results for the 345 diseases (170 ARD, 175 non-ARD) with available GWAS data (Supplementary Figure 18, Supplementary Table 16), while the male analysis fit Wald ratio results for 304 diseases (210 ARD, 94 non-ARD) with available GWAS data (Supplementary Figure 19, Supplementary Table 17). No results were phenome-wide significant.

### CETP – Tests for Group Effects

Permutation tests for disease-group effects in the both-sex analysis show a statistically significant (Benjamini-Hochberg q < 0.05) risk effect of increased CETP protein concentration on cardiovascular diseases (Figure 6). This implies CETP inhibition is protective of cardiovascular diseases in general which is also the intended indication for CETP inhibitor drugs. This finding for cardiovascular disease occurred at level two of the GBD disease hierarchy and is Benjamini-Hochberg significant (q < 0.05) against a comparison set of 18 disease groups. Permutation tests for level one and level three disease category effects as well as ageing phenome effects were all null (Supplementary Figures 20-22).

### Control Phenotypes – MR-PheWAS

We tested positive and negative outcome controls for *GLP1R* and CETP in the both-sex European population. A negative control should have no causal relationship with the exposure (*GLP1R* or CETP), and a positive control should have a causal relationship in the expected direction. We found that increased *GLP1R* expression had the expected significant protective effect on type 2 diabetes (E11) (p = 6.37 × 10^-10^) (Supplementary Table 3) and expected null effect on brown hair color (p = 0.31) (Supplementary Table 18). We found increased CETP concentration, the opposite effect of a CETP inhibitor drug, had the expected significant risk effect on high cholesterol (p = 6.25 × 10^-3^) (Supplementary Table 19) and null effect on brown hair color (p = 1.00) (Supplementary Table 20).

We also tested positive and negative exposure controls (smoking initiation and handedness) to confirm our MR-PheWAS pipeline produced expected phenome-wide results of known-risky and known-harmless exposures. Our negative control left-handedness returned expected null results for the both-sex European population after all exposure-outcome effects failed to pass robustness checks. We tested a second negative control, ambidextrousness, which returned expected null results in the European both-sex population, with no effect reaching phenome-wide significance (Supplementary Table 21, Supplementary Figure 23). Our positive control smoking initiation had the expected phenome-wide significant risk effect on lung cancer (C34) (q = 9.89 × 10^-5^) and chronic obstructive pulmonary disease (J44) (q = 4.49 × 10^-19^), with phenome-wide risk effects on a total of 50 diseases and protective effects on zero diseases in the European both-sex population (Supplementary Table 22, Supplementary Figures 24-25). Permutation tests for disease category effects did not reach statistical significance after multiple testing correction (Supplementary Figures 26-29). Results from MR-PheWAS of smoking initiation of East Asian both-sex and sex-specific analyses were either partial fits or non-fits due to data limitations, and uniformly without significant findings (Supplementary Figure 30, Supplementary Table 23). Additional detail on exposure control results is available in Supplementary Note 1.

## Discussion

We found significant causal relationships between *GLP1R* and ICD-10 coded diseases ranging across the human disease phenome for women, men, and both sexes. We identified 92 total effects and 66 novel effects of *GLP1R* in the both-sex phenome, underscoring the power of MR-PheWAS to identify opportunities and risks for drug repurposing.

Of particular interest was the largest risk effect of *GLP1R* on vitamin D deficiency (OR 2.72 per SD *GLP1R* expression), not found in the existing Mendelian randomization literature. Curiously, *GLP1R* levels appear to be positively correlated with vitamin D in human observational studies^37^, while intervention studies of *GLP1R* show risks of vitamin deficiencies^38^. Our findings align with intervention studies and suggest that GLP-1R agonists may raise risk of vitamin D deficiency. This finding is particularly relevant to populations vulnerable to vitamin D deficiency, such as women and people with a migration background living in the low-sun climates of Europe and North America^39^.

Secondly, we identify a risk effect of *GLP1R* on neonatal health. Our results show mixed effects of *GLP1R* on maternal diseases (37% risk effects), while we find a strong risk effect on neonatal diseases (75% risk effects). Of the two maternal disorders (O72 and O70) that *GLP1R* protects against with established clinically meaningful effect sizes (OR > 1.1), both are obstetric traumas where neonate birth weight is a risk factor. For postpartum hemorrhage (O72), a meta-analysis of 800 million women places high birth weight (>4kg) in the largest effect size category of risk factors^40^, and a second meta-analysis finds a monotonic relationship between birth weight and PPH risk with an odds ratio of 2.05 for birth weights above 4kg^41^. For perineal laceration during delivery (O70), the same meta-analysis finds an odds ratio of 1.90 for birth weights above 4kg.

We suggest our findings are compatible with *GLP1R* paradoxically protecting against obstetric trauma by inhibiting fetal growth, resulting in lower birth weights and reduced trauma during delivery. A similar effect is observed for cigarette smoking, which has been found to reduce obstetric trauma via inhibiting fetal growth^42^.

Mouse studies directly support the hypothesis that GLP-1 activity inhibits fetal growth, finding GLP-1-exposed pregnancies have reduced birth weights^8,9^ and increased risk of fetal death^9^. Emerging observational evidence in humans is mixed, with some studies finding a risk effect of GLP-1R agonists on maternal and birth complications^43^, while others find no effect^44^.

If GLP-1R agonism does present a risk to neonates, the anticipated increase in wider population usage and long washout period of current GLP-1R agonists may present a significant public health concern. In the United States, the median time to become aware of a pregnancy is 5.5 weeks, with 23% of pregnancies being recognized at or after 7 weeks^45^. The common GLP-1R agonist semaglutide (also known as Ozempic and Wegovy) has an effective clearance time of 5-6 weeks^10^. This suggests that GLP-1R-exposed pregnancies may have completed most or all of the first trimester under GLP-1R-exposure, even if drug use is discontinued immediately after the pregnancy is discovered. Our findings support the United States Food and Drug Administration current recommendation that pregnant women should not use GLP-1R agonists^46^, and raise a further concern about GLP-1R agonist use among women who may unknowingly be pregnant.

Emerging evidence suggests that GLP-1R agonists may function as anti-ageing drugs^6^. Despite the sizeable number of risk and protective effects of *GLP1R* expression identified in this study, we do not find evidence of an ageing-phenome specific effect of *GLP1R* for women, men and both sexes. This null finding for the effect of *GLP1R* expression on age-related disease despite many significant risk and protective findings may suggest that GLP-1R agonism is a double-edged sword, protecting against some age-related diseases while increasing the risk for others. Figure 5 shows that *GLP1R* expression has an expected within-ARD protective effect on diabetes and kidney diseases (5 protective, 0 risk), and a mixed-to-risky effect on musculoskeletal disorders (4 protective, 5 risk), with the risk concentrated on age-related bone degeneration such as Polyarthrosis (M15), Gonarthrosis (M17), and Spondylosis (M47).

Our findings show a stark contrast between the two molecular phenotypes of *GLP1R* expression and CETP concentration. While *GLP1R* expression affects most diseases we tested in the human phenome, CETP has only the narrow and anticipated effect on cardiovascular disease.

This suggests that unlike GLP-1R agonists, CETP inhibitors do not show potential for drug repurposing, nor high risk for side effects assuming well-targeted CETP inhibition. This is relevant to the ongoing research and development of CETP inhibitor drugs^12,13^, and supports the hypothesis that the hypertension risk observed in the first generation CETP inhibitor torcetrapib was due to an off-target effect rather than an unavoidable side-effect of CETP inhibition. We suggest this disparity between CETP and *GLP1R* underscores the ability of phenome-wide Mendelian randomization to distinguish between drug targets with high and low potential for repurposing.

Our study was limited by both data and methodological constraints, which may affect the interpretation of the outcomes. Our exposure of *GLP1R* gene expression levels is a related but distinct measure from the effect of GLP-1R agonist drugs. To address this concern, we used diabetes mellitus diagnosis (E11) as a positive control to check that *GLP1R* gene expression matched the known causal effect of GLP-1R agonism (Supplementary Note 1). This difference of exposure also complicates interpretation of log OR effect sizes. While our effect sizes are measured in standard deviations of circulating *GLP1R* transcripts among the untreated population, the dosage of GLP-1R agonist drugs is typically measured in circulating (unbound) levels of the drug, a distinct measure^47^. We note that these levels are commonly 20 to 30 times greater in magnitude than endogenous levels, suggesting the change in GLP-1 activity driven by drug exposure may be significantly larger than natural within-population variation^47^.

Our instruments for the sex-specific *GLP1R* sensitivity analyses come from a both-sex GWAS^33^ which may affect results if these instruments are uncalibrated in single-sex populations^20^ (Supplementary Discussion, *GLP1R* Instruments).

Due to the “absolute risk” scale used to define ICD-10 outcome phenotypes in the Neale Lab round 2 GWAS, it was not possible to compare risk effect sizes across disease in our sex-specific analyses. This limits the interpretation of sex-specific analyses.

Our outcome data comes from published summary statistics of the UK Biobank, without external cohort validation. To address this limitation, we completed sex-conserved validations where possible by treating the female UKB cohort as discovery and the male UKB cohort as a validation set (Supplementary Discussion, *GLP1R* Validation). This leaves the sex-specific findings, such as for maternal health, unvalidated.

We relaxed the p-value and quality control thresholds for instrument selection in the sex-specific analysis of CETP. As the sex-specific analyses of CETP did not produce any significant results, this preprocessing choice may not affect interpretation of the significant findings (Supplementary Discussion, CETP Instruments).

We identified a need for larger-than-typical statistical power for exposure GWAS, seemingly driven by the demanding nature of our quality control and robustness checks (Supplementary Discussion, Robustness Checks). Of 8 runs we completed with fewer than five exposure SNPs, only one contained significant results, while the other seven were either null (1), partial failures to fit (3) or total failures to fit (3).

We used permutation testing to detect disease-group-wide effects of exposures, but found the approach less sensitive than intended. This was compounded by multiple testing adjustment; of three nominally significant disease-group-wide effects of smoking initiation, none remained significant after multiple testing correction. We attribute this to limitations of permutation testing, which compares within-group effects to a null of outside-group effects, rather than no effect. This comparison tends to bias results toward the null, particularly when an exposure (such as *GLP1R*) has effects broadly distributed across the phenome rather than concentrated within a single disease group (as with CETP). We chose permutation testing due to data constraints (Supplementary Discussion, Permutation Testing).

## Methods

### 1. Overview of study design

We fit sex-specific (female, male and both sexes) phenome-wide Mendelian randomization analyses on CETP concentration and *GLP1R* expression levels, then tested for statistically significant effects of these exposures on disease groups and the phenome of age-related diseases (Figure 1). We used genetic instruments from sex-specific Genome-Wide Association Studies for all exposures excluding *GLP1R*, where we used both-sex exposure instruments under the assumption that genetic mutations affecting *GLP1R* expression levels are not meaningfully sex-specific^20^. The male-specific and female-specific outcome GWAS summary statistics we used are reported as absolute risk differences rather than log-odds ratios. This prevents comparison of MR effect sizes across diseases without referencing the baseline rate of disease; we therefore report p-values of male-specific and female-specific findings but not effect sizes. For both-sex analyses, we report both p-values and effect sizes. All statistical tests reported are two-tailed unless specified otherwise.

We used two types of positive and negative control phenotypes to separately check our exposure instruments and our MR-PheWAS pipeline. We used positive and negative outcome controls for *GLP1R* and CETP to confirm our instruments for these exposures capture the known effects and non-effects of *GLP1R* expression and CETP concentration. In addition, we used positive and negative exposure controls to confirm that our MR-PheWAS pipeline captured the expected effects of known risky exposures (smoking) and non-risky exposures (handedness) on the disease phenome (see Methods, Controls).

As a sensitivity check on the analysis of *GLP1R* in the both-sex population, we generated a “sex-conserved” list of significant *GLP1R* disease effects by treating the female MR-PheWAS of *GLP1R* as a discovery set, and validating significant findings using the results of the male MR-PheWAS with a Benjamini-Hochberg FDR q-value threshold of q < 0.05.

We applied permutation testing to test for significant differences of the beta coefficients generated by Mendelian randomization of age-related diseases and non-age-related diseases. Additionally, we applied permutation testing to Mendelian randomization beta coefficients grouped by GBD disease categories against outside-category beta coefficients to test for differential effects of the exposures on specific disease categories.

This study was designed with reference to the STROBE-MR guidelines for reporting Mendelian randomization analyses^31^ and Burgess et al.’s “Guidelines for performing Mendelian Randomization investigations,”^16^ with deviations adapted to accommodate phenome-wide Mendelian randomization. This study was not preregistered.

### 2. Data sources

#### Phenotypic hierarchy data

We used Global Burden of Disease survey^21^ (GBD) definitions as our primary disease ontology. GBD is a cross-country observational study that aggregates population-level disease diagnosis from 204 countries. These are contained in a hierarchy of increasing specificity^21^. The sex-specific ageing phenomes, containing lists of globally conserved age-related diseases, come from a forthcoming paper developing sex-specific ageing phenomes from GBD disease incidence data^30^.

#### Exposure data sources

The genomic data we used to proxy exposure to *GLP1R* expression and CETP concentration came from published GWASs, pQTL studies, and expression quantitative trait loci (eQTL) studies. Each study design contains associations between individual genetic mutations and observed clinical and molecular phenotypes.

The exposure data we used for *GLP1R* expression levels come from eQTLgen, a consortium of 37 blood-based expression quantitative trait loci cohorts not overlapping with the UK Biobank,

have a sample size (n) of 31,684, and have previously been used in Mendelian randomization studies of *GLP1R* expression^32,33,48^. The exposure data we used for both-sex CETP come from a GWAS of CETP concentration in whole blood with n = 4248 participants^35^. The exposure data we used for sex-specific CETP come from a sex-specific analysis of CETP using data from the Global Lipids Genetics Consortium multi-ancestry meta-analysis with a sample size (n) of 208^36,49^.

The exposure data we used for smoking among Europeans came from the Tobacco and Genetics (TAG) consortium^29^ and were accessed via the OpenGWAS catalog^50^. The TAG GWAS summary statistics used in this paper have an exact sample size (n) of 74,035.

TAG contains mostly European participants. Smoking data in the TAG consortium is self-reported^29^.

The exposure data we used for East Asian smoking comes from BioBank Japan, a single-cohort registry of participants with case status for one or more of 51 predetermined diseases ^27,28^. The BioBank Japan GWAS summary statistics used in this paper have an exact sample size (n) of 75798 (female), 89678 (male), and 165436 (both sexes). The cohort is overwhelmingly of Japanese ancestry. These data were accessed via OpenGWAS catalog^50^.

The exposure data we used for handedness come from the UK Biobank, and was published by Neale Lab as part of their UKB Round 2 GWAS project^22^. The UKB-based handedness GWAS summary statistics used in this paper have a sample size (n) of 337130 for both ambidextrousness and left-handedness. These data were calculated using only European ancestry UKB participants.

#### Outcome data sources

We used previously published genetic data for our phenome-wide outcomes from two sources of genome-wide association study summary statistics, both of which are based on GWAS in the UK Biobank (UKB)^51^. The UKB is a prospective cohort study containing around 500,000 participants from the UK population, including genomic data, multi-omic data, clinical data drawn from National Health Service (NHS) records and a participant questionnaire.

We used previously published summary statistics for sex-specific GWAS in UKB from the Neale Lab UKBB project round 2, an open-access project that calculated and hosts sex-specific GWAS summary statistics for 4,236 phenotypes in the UK Biobank^22^. We accessed the genomic variant manifest, sex-specific manifests, and sex-specific GWAS summary statistics from the Neale lab host server as part of an automated Mendelian randomization analysis pipeline using our R package “ardmr”^22^. All summary statistics downloaded from Neale Lab UKBB round 2 were calculated using the genomic reference build GRCh37 (hg19) and European-ancestry biobank participants only. Sex-specific Neale Lab Round 2 GWAS results used in this paper were calculated from an original sample (n) of 183,139 women and 154,060 men with available genotype data, with subsets taken where additional control participants did not add statistical power.

We used previously published both-sex summary statistics for ancestry-specific GWAS in UKB from the PanUKBB project, an open-access project associated with the Neale Lab that hosts GWAS summary statistics for 7,228 phenotypes across 6 continental ancestry groups^23^. We downloaded genomic files including variant manifests, GWAS summary statistic Tabix files, and selections of GWAS summary statistic files from the PanUKBB host server multiple times using our R package “ardmr”. All summary statistics downloaded from PanUKBB were calculated using the genomic reference build GRCh37 (hg19) and used combined male and female biobank participants. PanUKBB GWAS results used in this paper were calculated from an original sample (n) of 420,531 European-ancestry participants, with subsets taken where additional control participants did not add statistical power.

We accessed summary statistics for our outcome control phenotypes from the IEU OpenGWAS catalog^50^ except where these controls were already tested inside the MR-PheWAS pipeline. The outcome data we used for the CETP positive outcome control high cholesterol diagnosis comes from a GWAS of age-related diseases in the predominantly European-ancestry UKB biobank^26^ using a multi-ancestry framework. The outcome data we used for the *GLP1R* positive control type 2 diabetes (E11) was measured in PanUKBB and was tested automatically through the MR-PheWAS pipeline^23^. The outcome data for our *GLP1R* and CETP negative outcome control, naturally dark brown hair, comes from a GWAS in the UK Biobank^25^. The phenotype for brown hair color was defined as self-reported “dark brown hair color (natural, before greying).”

### 3. Mendelian Randomization PheWAS

#### Overview of Mendelian randomization assumptions

Mendelian randomization is a statistical method for estimating the causal effect of an exposure on an outcome using genetic instrumental variables^17^. MR leverages the recombination of the two parental genomes during conception as a source of random variation, and uses instrumental variable methods to estimate the effect of these randomly assigned alleles^17^. The assumptions of Mendelian randomization are available in Supplementary Methods, Mendelian Randomization Assumptions^17^.

### 3.1 Selection of genetic instruments

#### Exposure instruments

We identified instruments for our exposures from published genome-wide association studies, protein-quantitative trait loci studies, expression quantitative trait loci studies, and through the openGWAS catalog. When drawing instruments from prior studies, we followed the preprocessing standard of the relevant publications. Our instruments for GLP1R come from eQTLgen, following the preprocessing pipeline set by a GLP1R Mendelian randomization study from Sun et. al^32^, since used by several other Mendelian Randomization studies of GLP1R^52–56^; eQTLs in a 1Mb window of the GLP were filtered for minor allele frequency greater than 1%, significant association with GLP1R expression at P < 5 × 10⁻⁸, and association strength test at F > 10.

Our both-sex instruments for CETP come from a GWAS of CETP^35^; SNPs were selected based on genome-wide significant (P < 5 × 10⁻⁸) association with CETP. All passing SNPs were cis-pQTLS of CETP. Independence was established by performing stepwise conditional and joint association analysis in GCTA (genome-wide complex trait analysis) v1.24.4 to run conditional and joint analyses with a stepwise selection procedure to identify variants independently associated. Separately to Blauw et al, we applied filtering for strength of association at a threshold of F > 10 to all passing SNPs to address weak instrument bias risk in our MR study.

Our sex-specific instrument for CETP come from a sex-stratified lipid GWAS meta-analysis with follow-up colocalization by Kanoni et al^36^. The selected variant (rs247617) is a cis-eQTL of the *CETP* gene in adipose tissue, as measured in the Gene-Tissue Expression Project^57^, identified by Kanoni et al as having a sex-divergent effect on blood lipids. This analysis used the Benjamini-Hochberg adjusted significance threshold of q < 0.05 to establish sex-specific eQTL status with CETP, notably lower than the standard genome-wide significance threshold for MR.

Our instruments from the seven outcome control analyses were accessed via the openGWAS catalog, and were selected using standard MR preprocessing steps^16^. We selected exposure instruments from exposure GWAS using the genome-wide significance p-value threshold (p = 5 × 10^−8^).

Once instruments were selected, we performed linkage disequilibrium (LD) clumping with threshold 𝑟^2^ = 0.001 across a 10,000 kb window, and dropped SNPs with 𝐹 < 10. We then applied pleiotropy screening on an as-needed basis for highly confounded (behavioral) phenotypes using the PheWAS function in the TwoSampleMR package (Table 1) (Supplementary Methods, Pleiotropy Screening).

#### Outcome Instruments

We obtained instruments for the outcomes in the MR-PheWAS of each ageing phenome using publicly available GWAS summary statistics of the UKB^22,23^.

We started the outcome-instrument gathering process by mapping diseases from the GBD to ICD-10 code GWASs with available summary statistics in the PanUKBB or Neale Lab round 2 UKB GWAS using a GBD-published mapping of diseases-to-codes. Then, we constructed an outcome plan that listed all outcome phenotypes to be analyzed in that MR-PheWAS run, along with their status as age-related diseases or not (ARD selection status), GBD category values to allow for permutation tests of ARD and category enrichment, and GWAS summary statistics for all available outcome phenotypes We then performed a pre-MR harmonization on the exposure and outcome SNPs. We aligned the effect allele and non-effect allele between exposure and outcome to prevent sign flips. We then resolved palindromic alleles using effect allele frequencies. Finally, we filtered and dropped allele mismatches.

#### Controls

We used the standard MR practice of testing our instruments for *GLP1R* and CETP against positive and negative outcome control phenotypes^16^. This step was intended to confirm that MR using these instruments identify presumed-true causal relationships of our exposures (positive control) and fail to identify presumed-false causal relationships of our exposures (negative control). For *GLP1R* exposure, we used type 2 diabetes^23^ (E11) diagnosis as measured in the European both-sex population as our positive outcome control phenotype, as GLP-1R agonists were originally approved to treat type 2 diabetes. For our CETP exposure, we used high cholesterol^26^ as measured in the European both-sex population as our positive outcome control phenotype, as the CETP inhibitor drug Obicetrapib is under review to treat high cholesterol^13^. For both *GLP1R* and CETP, we used brown hair color^25^ in the European both-sex population as our negative outcome control phenotype.

In addition to using positive and negative outcome controls to validate our exposure instruments, we also tested our MR-PheWAS pipeline with positive and negative exposure controls. This was intended to confirm expected behavior of our MR-PheWAS pipeline under known-risky and known-harmless exposures, including across multiple sexes and ancestry groups. For positive exposure controls, we used smoking initiation in the European both-sex population^29^, smoking initiation in the East Asian both-sex population^27,28^, and smoking initiation in the East Asian men and women^27,28^ separately. These GWAS had no sample overlap with our UKB-based outcome GWASs, which we deemed necessary for positive exposure controls. We predicted smoking initiation would be a significant risk factor for “C34 Malignant neoplasm of bronchus and lung” and “J44 Other chronic obstructive pulmonary disease,” and not protective of any disease from the level-2 GBD categories “Chronic respiratory diseases” or “Cardiovascular diseases”. For negative exposure controls, we used left-handedness and ambidextrousness, both measured in the European both-sex population^22,27,29^. We predicted left-handedness and ambidextrousness would not have phenome-wide significant effects on any disease. The instruments for these two phenotypes come from GWASs in the UK Biobank, indicating nearly complete sample overlap with our phenome-wide outcome data. We deemed this acceptable for a negative but not positive control phenotype due to overlap driving results away from the null^58^. We predicted handedness would not be significantly associated with any disease.

### 3.2 MR-PheWAS Implementation

#### Primary estimators

After the exposure and outcome SNPs were quality controlled and harmonized, we conducted an MR-PheWAS by fitting a primary MR estimator of the exposure to each mapped outcome in the GBD-defined phenome. When conducting the MR-PheWAS, we used the Wald ratio as the primary estimator for exposure phenotypes with only one instrument surviving quality control^17^. The Wald ratio for exposure SNP J is shown in equation (1) and describes the estimated regression coefficient of SNP J on the outcome 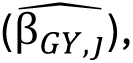 scaled by the estimated regression coefficient of the exposure SNP on the exposure 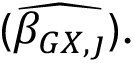 The variance of the Wald ratio used to calculate confidence intervals shown in equation (2) is estimated using the delta method, which returns a linear approximation of the true variance^59^.

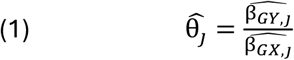

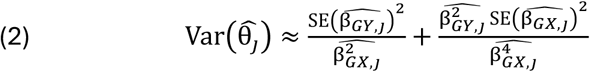

We used the inverse variance weighted (IVW) method as the primary estimator for exposure phenotypes with more than one instrument surviving quality control. IVW estimates the true effect of the exposure on the outcome using equation (3). Equations (3) and (4) show a precision-weighted estimate of the Wald ratios of K exposure SNPs, where the weight of SNP j is determined using the estimated variance of the Wald ratio (4). The confidence intervals for 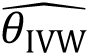 in equation (3) are estimated using the variance formula shown in equation (5).

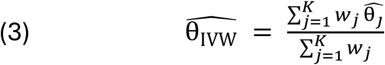

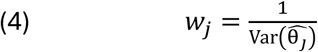

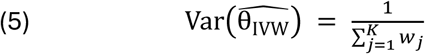

#### Robustness Checks

Due to the hundreds of MR estimators fit in a single MR-PheWAS of a single population, it was infeasible to manually apply and interpret robustness checks as is typical for single-outcome MR^16^. Instead, we employed an automated robustness check procedure based on prior phenome-wide MR publications^15,60,61^. We applied eight robustness checks to each exposure-outcome MR where the number of exposure SNPs (i.e., instrumental variables) was greater than one, then dropped outcomes from the MR-PheWAS that passed less than a predetermined threshold number of tests (Supplementary Table 1). We chose our robustness checks using guidance from MR methods papers that recommend multiple robustness checks with varied assumptions to reduce the risk of hidden assumption violations, including at least one weighted median and one weighted mode robustness check^16,24^. Our robustness checks were as follows:

1. We calculated MR-Egger, which relaxes the exclusion-restriction assumption, allowing for horizontal pleiotropy but requiring that the pleiotropic effects are independent of the instrument-exposure association strength. This is also referred to as the InSIDE assumption^16^. We coded sign disagreement of the MR-Egger slope with the primary IVW slope as a failure.
2. We subjected the intercept of the MR-Egger regression to a hypothesis test against a null of 𝐻_0_: intercept = 0. If we found an intercept not equal to zero with a p-value < 0.05, we coded the test as a failure.
3. We calculated Cochran’s Q, which tests for heterogeneity of instrument effects^61^. If we found heterogeneity with a p-value < 0.05, the test was recorded as a failure.
4. We calculated 𝐼^2^as a function of Cochran’s Q^62^, which estimates the proportion of observed heterogeneity that is attributed to true heterogeneity of instrument effect, rather than sampling error. If we found 𝐼^2^> 50%, the test was recorded as a failure.
5. We calculated weighted median of the Wald ratios and checked for sign disagreement of the slope with the IVW estimate^16^. Sign disagreements were recorded as a failure.
6. We calculated the weighted mode of the Wald ratios and checked for sign disagreement of the slope with the IVW estimate^16^. Sign disagreements were recorded as a failure.
7. We calculated Steiger directionality tests for each instrument to check whether the observed strength of the associations indicated that the instrument had a large impact on the outcome rather than the exposure, which would suggest pleiotropy or reverse-causation^15^. If more than 30% of the instruments failed the Steiger directionality test, we recorded a robustness check failure.
8. We calculated a leave-one-out (LOO) analysis of the IVW estimate^63^. We then checked for sign flips of the slope under all the LOO folds, which would suggest heterogeneity of the effect between instruments. If we discovered a sign flip in any fold, the test was recorded as a failure.

We did not apply this robustness check procedure to exposures with only a single instrumental variable surviving initial quality control (QC), as all checks except for Steiger directionality could not be implemented. Instead, outcomes in single-instrument MR-PheWAS were dropped only if their Wald ratios were undefined. Due to the reduced confidence in the phenome-wide validity of these MR-PheWAS’, we reported the individual MR results but did not conduct tests for a phenome-wide effect on age-related disease, or on any disease category.

After filtering exposure-outcome pairs that failed the MR sensitivity analysis check, we applied Benjamini-Hochberg false discovery rate adjustment to the p-values of surviving outcomes. We reported outcomes as significant if their β coefficient differed from zero with q < 0.05 after multiple testing correction.

### 4. Phenome-wide effects on age-related disease

We used permutation testing to identify an aggregated effect of our exposures on the sex-specific ageing phenomes, which are a subset of the full phenome explored in the MR-PheWAS. For the permutation tests, we constructed signed disease scores 𝑠_𝑖_ for each surviving MR outcome result by multiplying the negative 𝑙𝑜𝑔_10_ p-value of the IVW beta coefficient 𝑝_IVW,𝑖_ by sign(𝛽_IVW,𝑖_), as shown in equation (6). This signed disease score is intended to capture both confidence and direction of an effect for that disease. Then, we constructed the observed test statistic 𝑇_obs_ that captures the precision-weighted mean of the signed disease scores across diseases in the ageing phenome shown in equation (7). As shown in equations (8) and (9), 𝑦_𝑖_ is ARD selection status and 𝑤_𝑖_ is weight given by the inverse of the squared standard error of the IVW estimate for that disease.

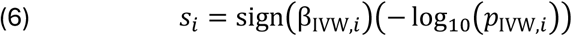

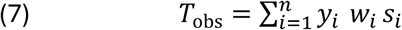

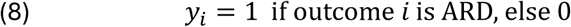

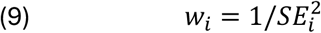

We then conducted permutation testing on 𝑇_obs_ against a null hypothesis that ARD selection status (𝑦_𝑖_) is irrelevant to the distribution of 𝑇. We did this by shuffling the ARD selection status labels over 𝑏 (=10,000) permutations. From this we constructed an estimate of the mean and standard deviation of 𝑇 under the null hypothesis, shown in equation (10) and (11). Using equations (10) and (11), we conducted our hypothesis test on 𝑇_obs_, yielding a signed enrichment score (SES) shown in equation (12) and a p-value for SES shown in equation (13). These represent the standardized distance of the observed ARD disease scores 𝑇_obs_ from the null hypothesis, and the degree of confidence that 𝑇_obs_ ≠ 𝑇_null_.

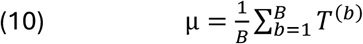

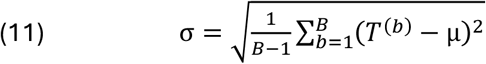

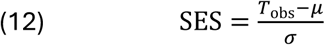

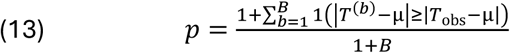

#### GBD Category Testing

We also applied the same permutation testing methods to examine whether exposures had a category-wide effect on diseases by GBD category. We conducted category-wide permutation tests for each category in GBD hierarchy levels one, two and three. In this case, 𝑇_obs_ was defined as the precision-weighted sum of the signed disease scores included in that category, or for the children of diseases included in that category. For example, the level two disease “neoplasms” has a level one parent “non-communicable disease” and level three child “leukemia.”

Accordingly, the permutation test of “neoplasms” included the beta coefficient for the child “leukemia” but not for the parent “non-communicable disease.” Due to the risk of multiple testing, we applied Benjamini-Hochberg FDR adjustment within each hierarchy. Where all results were null, we repeated the analysis, relaxing the multiple testing correction for the both-sex analyses. We did not repeat analyses with relaxed multiple testing correction for female-specific and male-specific analyses.

#### Maternal and Neonatal Diseases

We wished to analyze the effect of *GLP1R* on maternal diseases, and separately the effect of *GLP1R* on developing fetuses. We identified the GBD category “Maternal and neonatal disorders” as a close match for this aim, but noted that many ICD-10 codes (15 of 33) mapped to this category referred to billable but non-diagnostic maternal medical events, such as screening (Z36 Antenatal screening), care (Z39 Postpartum care and examination), non-pathological medical outcomes (O80 Single spontaneous delivery), or procedures (O04 Medical abortion).

To capture disease diagnosis risk, we excluded the 15 ICD-10 codes not referring to diagnoses of maternal or neonatal pathologies, retaining 18 diseases. We then separated the remaining diseases into the categories “neonatal diseases” and “maternal diseases,” which we drew from obstetrics literature^64^, using ICD-10 code definitions from the World Health Organization 2019 issue of the International Statistical Classification of Diseases and Related Health Problems 10th Revision. The category “neonatal diseases” captures health risks to either the mother or fetus originating from the fetus or from pathological fetal development, such as “O68 Labor and delivery complicated by fetal stress.” The category “Maternal diseases” captures health risks to either the mother or fetus not originating from the fetus, such as “O21 Excessive vomiting in pregnancy.” We assigned diseases originating from the umbilical cord as belonging to both the mother and the neonate as the umbilical cord has no clear owner after birth.

## Data Availability

The data used in this study are openly available from the Global Burden of Disease survey^21^, eQTLgen^32,33^ for *GLP1R* expression, a GWAS of Netherlands Epidemiology of Obesity cohort for both-sex CETP concentration^35^ and sex-specific CETP concentration from the Global Lipids Genetics Consortium multi-ancestry meta-analysis^36,49^. Exposure data for smoking comes from the Tobacco and Genetics (TAG) consortium^29^ for European ancestry, and from BioBank Japan ^27,28^ for East Asian ancestry, both accessed via OpenGWAS catalog^50^. The phenome-wide GWAS summary statistic data come from the Neale Lab UKBB GWAS round 2^22^ and the PanUKBB project for 7,228 phenotypes across 6 continental ancestry groups^23^. Data for ambidextrousness and left-handedness additionally come from Neale Lab UKBB GWAS round 2^22^. Both PanUKBB and Neale Lab GWAS round 2 are based on data from the UK Biobank^51^.

## Code Availability

The scripts used to perform all coded analyses and generate all figures in this analysis are contained in the supplement and available on a GitHub repo^65^. Main analysis including all MR-PheWAS and permutation tests were conducted with our R package “ardmr,” a tool for conducting easy testing of exposures on the full human disease phenome, as well as the sex-specific age-related disease phenomes using the pipeline described in Methods. Full examples and documentation are available in the readme file of ardmr hosted on the package GitHub repo^66^.

Outside of the ardmr package, we performed basic data cleaning as described in Methods using R v4.4.3 and the packages readr v2.1.5, dplyr v1.1.4, stringr v1.5.1, tidyr v1.3.1, ggplot2 v3.5.2, forcats v1.0.0, grid v4.4.3, ggrepel v0.9.6, scales v1.4.0, purrr v1.1.0, tibble v3.3.0.

## Data Availability

All data produced in the present study are available upon reasonable request to the authors.

https://pan.ukbb.broadinstitute.org/

https://www.nealelab.is/uk-biobank

https://opengwas.io/datasets/

https://github.com/Robert-Henry-Campbell/GLP1R_CETP_scripts

https://github.com/Robert-Henry-Campbell/ARD-Mendelian-Randomization

## Acknowledgements

R.H.C. is supported by the Leverhulme Trust Biopsychosocial Doctoral Training Programme (DS-2020-053), granted to M.C.M. M.C.M. is supported by ESSGN (HORIZON-MSCA-DN-2021 [101073237]), ESRC/UKRI Connecting Generations (ES/W002116/1) and Einstein Foundation Berlin (EZ-2019-555-2). R.H.C. and M.C.M are supported by the Leverhulme Trust Large Centre Grant LCDS (RC-2018-003).

We thank Benjamin Woolf for his expert advice on the strengths and limitations of Mendelian Randomization, and for his help selecting sensitivity and control analyses.

## Author Contributions

R.H.C. conceptualized the project, led the study design and data analysis, created all display items, the R package and drafted the initial manuscript. M.C.M. contributed to drafting the initial manuscript, revision and secured funding and supervised the project. All authors reviewed the manuscript for accuracy, edited subsequent revisions of the draft, and approved the final version.

