## Supplemental Text for "58 risk and 34 protective effects of *GLP1R* on disease phenome and adverse neonatal health"

### Supplement to “*GLP1R* expression protects against 58 diseases but raises risk for 34 diseases and neonatal health”

### Supplementary Notes

**Supplementary Note 1: Control Phenotypes**

We used exposure control phenotypes to test our MR-PheWAS pipeline, as well as outcome control phenotypes to test the behavior of our *GLP1R* and CETP exposure instruments.

We tested the effect of smoking initiation in the European both-sex population both with and without pleiotropy filtering exposure SNPs on obesity and education-related outcomes. This was intended as a sensitivity analysis of our positive exposure control of smoking. When including pleiotropy filtering, only 154 of 396 outcomes had available data for the single SNP instrument (Table 1, Supplementary Table 1). No results reached significance at Benjamini-Hochberg q < 0.05. Permutation testing was not performed because robustness checks were not applied due to the singular SNP exposure instrument. To increase the power of our positive control instruments, we abandoned the pleiotropy filtering on educational and adiposity related outcomes and reran the MR-PheWAS pipeline on smoking initiation (Table 1). In this relaxed run, reported in Results, 92 exposure SNPs passed quality control, and 300 disease outcomes tested passed robustness checks. Smoking initiation appeared as a phenome-wide significant risk factor for 50 diseases, and protective of no diseases (Supplementary Table 22, Supplementary Figures 24,25). The two strongest hits, which differed by ~20 orders of magnitude, were “Mental and behavioral disorders due to use of tobacco” (q = 6.18 × 10^-41^, ICD-10 code = F17) and “Other chronic obstructive pulmonary disease” (q = 4.49 × 10^-19^, ICD-10 code = J44).

Naïve grouping by category shows a risk effect of smoking on nine diseases from the “chronic respiratory disease” category (GBD cause level 2), as well as four mental and substance use disorders including alcoholism (F10) and depressive episodes (F32). Permutation tests for disease-group effects on age-related disease and GBD disease categories were directionally expected but did not reach statistical significance after multiple testing correction (Supplementary Figures 26-29). The strongest GBD group findings prior to multiple testing correction were for “substance use disorders” (p = 0.005), “chronic obstructive pulmonary disease” (p = 0.022), and “atrial fibrillation and flutter” (p = 0.037).

We additionally ran MR-PheWAS of smoking initiation in the East Asian both-sex population. We did not include pleiotropy screening due to concerns about loss of statistical power. For the East Asian both-sex population, 20 phenotypes (12 ARD, 8 non-ARD) passed robustness checks. Of these, none were significant. Permutation analysis was null at all levels.

We attempted sex-specific MR-PheWAS of smoking initiation in the East Asian female and male populations. No instruments survived exposure quality control, terminating these runs before results.

For the negative control phenotype of left-handedness in the European both-sex population, two exposure SNPs passed quality control, and none of the 396 disease outcomes tested (143 ARD and 172 non-ARD) passed MR robustness checks. For the negative control of ambidextrousness, only one exposure SNP passed quality control, and 118 outcomes (68 ARD and 50 non-ARD) had available outcome SNP data. We did not apply the robustness check pipeline as it is undefined on single-instrument exposures. No surviving results were significant at Benjamini-Hochberg adjusted q < 0.05 (Supplementary Table 21, Supplementary Figure 23). We did not conduct permutation testing on the left-handed phenotype as it had no robust results, or on the ambidextrous phenotype as it could not be robustness checked.

### Supplementary Discussion

***GLP1R* Instruments**

Our main analysis of *GLP1R* used exposure instruments derived from eQTLgen, an international consortium of gene expression quantitative trait loci from both-sex human blood^1^. We used these both-sex instruments in all analyses of *GLP1R*, including for the male and female specific runs. This data limitation presents a potential concern for the sex-specific analyses if the both-sex instruments are uncalibrated for predicting *GLP1R* expression levels in a single-sex population. To mitigate this concern, we searched the literature for and did not find evidence that *GLP1R* has meaningfully sex-specific QTLs. This finding concurs with the broader set of findings that blood QTLs generally show little evidence of sex-specificity^2^.

***GLP1R* Validation**

To address our lack of a validation cohort for our *GLP1R* findings, we performed a within-UKB biobank sensitivity analysis by treating the MR-PheWAS in women as discovery, and the MR-PheWAS in men as validation. This technique was made possible by the sex-specific GWAS published by the Neale Lab round 2 GWAS of the UK Biobank^3^, which allows for partitioning the participants without access to the individual level data. While this validation method cannot validate sex-specific effects, we suggest it robustly confirms the sex-conserved nature of effects observed in the both-sex population. Accordingly, sex-specific findings from the MR-PheWAS of *GLP1R* should be treated as unvalidated.

**CETP Instruments**

Data limitations forced a trade-off between using both-sex instruments for sex-specific analysis as we did with *GLP1R*, or alternatively using sex-specific instrument(s) selected with lower-than-standard selection criteria. We opted for the latter option, marking deviations from standard best practice with “*” and “**” in Table 1 and Supplementary Table 1. Our sex-specific analyses of CETP did not produce any significant results, thus we do not believe this preprocessing choice undermines the significant findings of this paper.

**Robustness Checks**

Our robustness check procedure commonly left zero or only a few surviving outcomes, which reduced the “phenome-wide” nature of the findings. Due to the hundreds of MR estimators fit in a single MR-PheWAS, it was infeasible to manually interpret each set of robustness checks. To address this, we implemented a broader array of robustness checks than would be used for a single-outcome MR with the intention of capturing most sources of assumption violations across phenotypes^4^, meeting the standard set by other phenome-wide Mendelian randomization studies^5–7^. However, we hypothesize that the automated nature of these tests makes them less discerning than manually interpreted tests, and forces a larger sensitivity-specificity trade-off. We chose to prioritize specificity, which resulted in null findings or complete or partial failures-to-fit tests for 9 of the 14 MR-PheWAS we ran. We suggest the importance of further development of automated robustness checks in phenome-wide Mendelian randomization.

**Permutation Testing**

Permutation testing was chosen as a method for group effect testing due to data constraints. The preferred approach would have been a t-test of the mean within-group MR effect against zero, but this requires estimating the covariance structure of MR beta coefficients. Assuming zero covariance was considered inappropriate, as it would systematically bias results away from the null. The permutation test instead makes weaker assumptions: under the null, disease scores and weights are assumed exchangeable between ARD and non-ARD labels. While the sign of MR effects should be randomly distributed around zero if MR assumptions hold, the associated p-values and standard errors may not be exchangeable. This assumption may be violated if ARD and non-ARD outcomes differ in GWAS power or in their likelihood of passing MR robustness checks, potentially biasing results toward or away from the null.

Future work could address limitations to this approach by permuting only effect signs, ignoring effect size and precision, or by using individual-level data to estimate covariance structures or to test category-wide effects directly through aggregated GWAS outcomes. This first option would reduce power while reducing bias, while the second, if possible, may increase power while reducing bias.

### Supplementary Methods

**Mendelian Randomization Assumptions**

Mendelian randomization critically depends on three assumptions to be valid^8^.

1. Relevance: all genetic instruments used must impact the exposure through some causal pathway. This assumption is typically satisfied by selecting instruments using the results of GWAS on the exposure of interest. This method identifies instruments which are significantly associated with the MR exposure phenotype.
2. Independence: all instruments must be independent of confounders. This criterion is credibly met for within-family MR designs, where genetic confounding is automatically adjusted for. However, most implementations of MR, including ours, use “two sample MR,” which creates a risk of non-random assignment of genetic instruments under assortative mating or population stratification^8^. However, non-family-based MR studies typically reduce the risk of confounding from the standard set by traditional epidemiological studies.
3. Exclusion restriction: all instruments must impact the outcome only through their effect on the exposure. Horizontal pleiotropy, where an instrument impacts the outcome through a causal pathway that does not include the exposure, violates this assumption. This risk is typically adjusted for by removing instruments strongly associated with confounders of the intended outcome, also called pleiotropy screening.

**Pleiotropy Screening**

Due to the phenome-wide nature of this MR study, we determined it would bias results to apply pleiotropy screening on outcomes that appear as phenome-wide endpoints, or on intermediate phenotypes such as lipid levels that are causally connected to diseases that appear as outcomes. We intended to take a hypothesis-free approach to the effects of each exposure and therefore did not want to rule out valid causal pathways leading to at least one disease present as an outcome phenotype. However, we assumed pleiotropy screening was useful on an as-needed basis to separate the effects of highly correlated behavioral phenotypes not likely to appear as ICD-10-coded endpoints. For this reason, we performed MR-PheWAS of our positive exposure control of smoking initiation both with and without pleiotropy screening for educational and adiposity phenotypes.

We used regex to match phenotype strings to multiple PheWAS hits with uncertain phrasing, for example using “educat” to capture both “educational attainment”, and “years of education” (Table 1). SNPs with genome-wide significant PheWAS that matched to a specified regex string were dropped.

**Outcome Plan**

The outcome plan contained all GWAS-linked diseases in the Global Burden of Disease survey, but diseases differed in ARD selection status depending on the sex of the population analyzed. ARD selection status is defined by sex-specific ageing phenomes^9^.

In cases of one-to-many mapping, where a single GBD disease was mapped to multiple ICD-10 codes, we preserved all codes as their own outcomes, and they inherited the same ARD status as their parent. We resolved cases of many-to-one mappings to the single ICD-10 code outcome. We decided ARD status for this surviving ICD-10 code outcome by majority vote of the ARD status on the multiple diseases mapped to it. We decided ties in favor of positive ARD status. We defined GBD hierarchy group values uninformatively from the first duplicate mapped disease with the majority vote ARD status.

Next, we gathered outcome SNPs for every GWAS into the outcome plan. We implemented this differently depending on whether the population studied included both male and female participants, or only a single sex. In the case of a both-sex population of any ancestry, we gathered outcome SNPs for each outcome phenotype using a Tabix interaction with the GWAS summary statistic files hosted on a PanUKBB-owned AWS cloud. Tabix is an indexing tool for genomic files that allows users to query summary statistics files by genomic coordinates, allowing for rapid retrieval^10^. We used Tabix to query only the outcome SNPs needed from the PanUKBB GWASs, allowing for streaming of the outcome SNPs in real time. Once a run had been completed, we saved a lightweight R dataset file containing only the streamed outcome SNPs in a local cache to save bandwidth for repeated runs.

In the case of a single-sex MR-PheWAS run, we extracted SNPs for a given outcome from a locally cached copy of the entire GWAS summary statistics for that outcome. If these files did not exist for all outcomes, we downloaded the missing files from a Neale Lab-owned AWS cloud containing their published GWAS summary statistics^3^. Once all outcome GWAS were available, we pulled the outcome SNPs required for the run with a non-Tabix search of each GWAS summary statistics file.

**Prior literature search**

For both CETP and *GLP1R*, we searched for prior MR and MR-PheWAS studies of the form shown in Equation 14 with results sorted by date. Searches were performed on January 19^th^ 2026.

(14)	**(TARGET[Title/Abstract]) AND (Mendelian Randomization[Title/Abstract])** (TARGET ∈ {CETP, GLP1R, GLP-1R, glucagon-like peptide 1 receptor})

**Literature Search for Novel Findings**

For each significant *GLP1R* disease finding in the both-sex analysis, we searched PubMed for terms “Mendelian randomization GLP-1R <disease>” in all search fields. We specified <disease> first as the ICD-10 coded name without the code, then using variations, simplifications and synonyms if no results were found. If no results were found through PubMed, we fell back to the Perplexity AI search engine, asking, “can you find a Mendelian randomization study of GLP-1R on <disease>?^11^” If no relevant results were found in either the PubMed searches or Perplexity searches, we recorded the finding as a novel Mendelian randomization finding.

11.	*Perplexity*. https://www.perplexity.ai/.
