## Supplemental Figures for "58 risk and 34 protective effects of *GLP1R* on disease phenome and adverse neonatal health"

Supplementary Figure 1. Phenome-wide MR results for GLP1R, European female population.

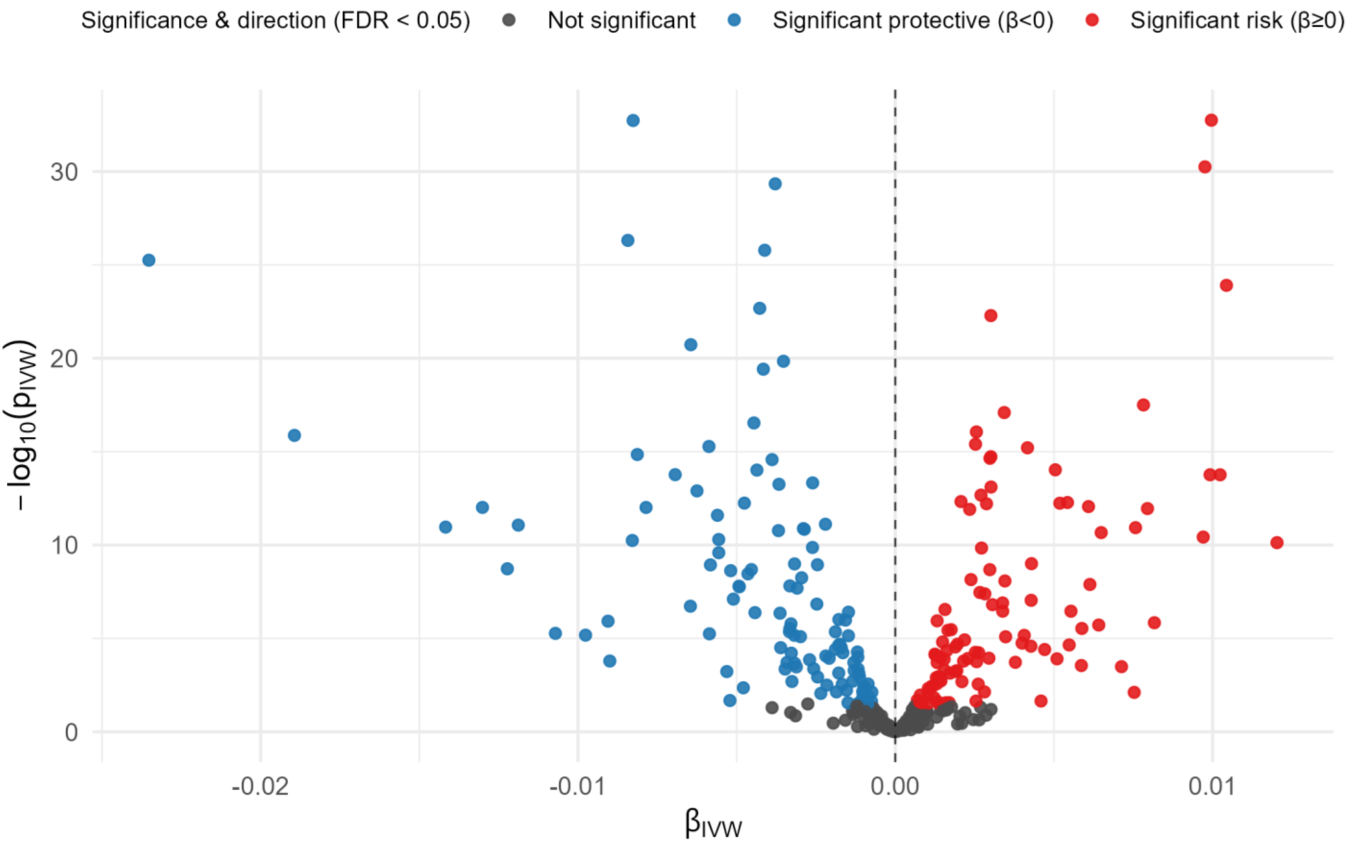


**Supplementary Figure 1.** Volcano plot results of phenome-wide Mendelian randomization of GLP-1R expression in female European population. “MR” refers to Mendelian Randomization testing. “GLP-1R expression” refers to genetically proxied GLP-1R expression. “FDR” refers to Benjamini-Hochberg false discovery rate correction. “βivw” refers to the causal effect of one standard deviation increase of GLP-1R expression on log-odds ratio of disease diagnosis as estimated using the inverse variance weighted method. The dotted line refers to a log-odds ratio of zero, implying no effect change in disease risk resulting from increased GLP-1R expression. Disease labels are applied to selected novel Mendelian randomization findings with selective labelling. Red dots imply increased GLP-1R expression raises risk for the named disease outcome, blue dots imply increased GLP-1R expression lowers risk, and grey dots represent null findings. Statistical significance is defined as $q < 0.05$ after Benjamini-Hochberg false discovery rate adjustment.

Supplementary Figure 2. Phenome-wide MR results for GLP1R, European male population.

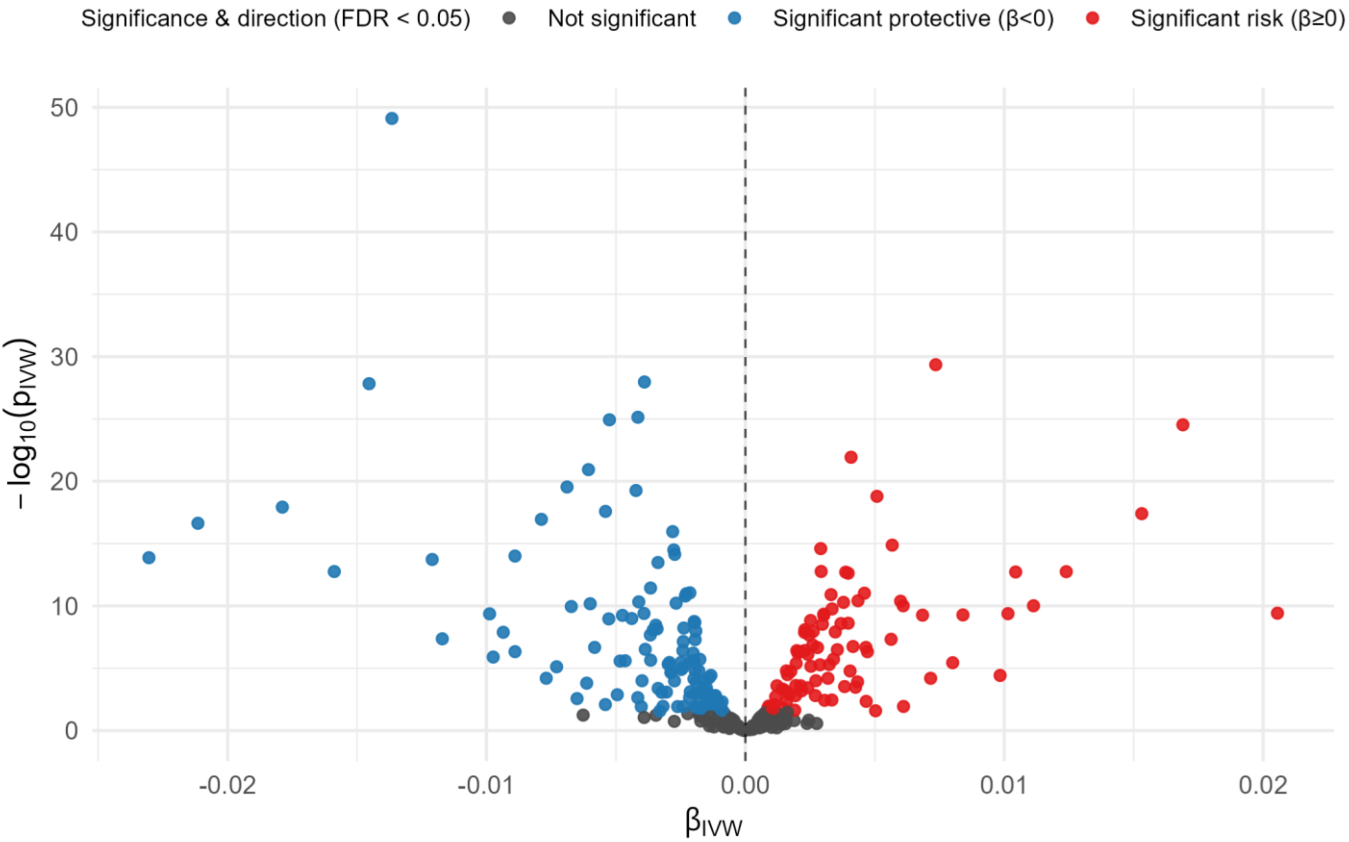


**Supplementary Figure 2.** Volcano plot results of phenome-wide Mendelian randomization of GLP-1R expression in male European population. "MR" refers to Mendelian Randomization testing. "GLP-1R expression" refers to genetically proxied GLP-1R expression. "FDR" refers to Benjamini-Hochberg false discovery rate correction. "βivw" refers to the causal effect of one standard deviation increase of GLP-1R expression on log-odds ratio of disease diagnosis as estimated using the inverse variance weighted method. The dotted line refers to a log-odds ratio of zero, implying no effect change in disease risk resulting from increased GLP-1R expression. Disease labels are applied to selected novel Mendelian randomization findings with selective labelling. Red dots imply increased GLP-1R expression raises risk for the named disease outcome, blue dots imply increased GLP-1R expression lowers risk, and grey dots represent null findings. Statistical significance is defined as $q < 0.05$ after Benjamini-Hochberg false discovery rate adjustment.

Supplementary Figure 3. ARD-wide MR results for GLP1R, European both-sex population.

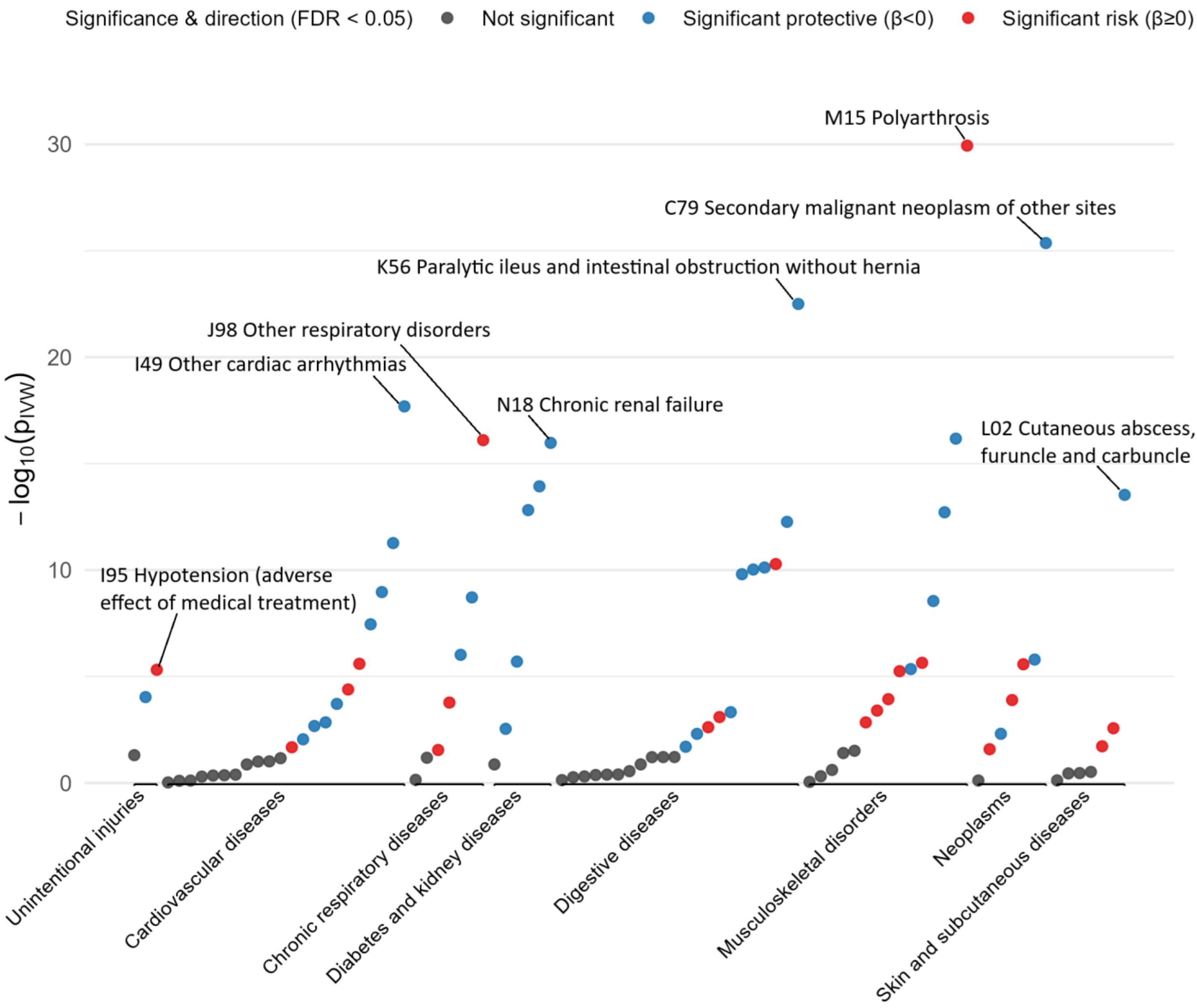


**Supplementary Figure 3.** Manhattan plot of GLP-1R expression effects on the phenome of age-related diseases in the both-sex European population. Diseases are grouped by Global Burden of Disease category (level 2). Disease labels are applied to the lowest p-value effect in each category, and to significantly affected diseases in the most risk-affected and protective-affected categories. Red dots imply increased GLP-1R expression raises risk for the named disease outcome, blue dots imply increased GLP-1R expression lowers risk, and grey dots represent null findings. Statistical significance is defined as q < 0.05 after Benjamini-Hochberg false discovery rate adjustment. -log10(Pivw) represents the log-scaled p-value measuring the statistical significance of the measured effect.

Supplementary Figure 4. ARD-wide MR results for GLP1R, European female population.

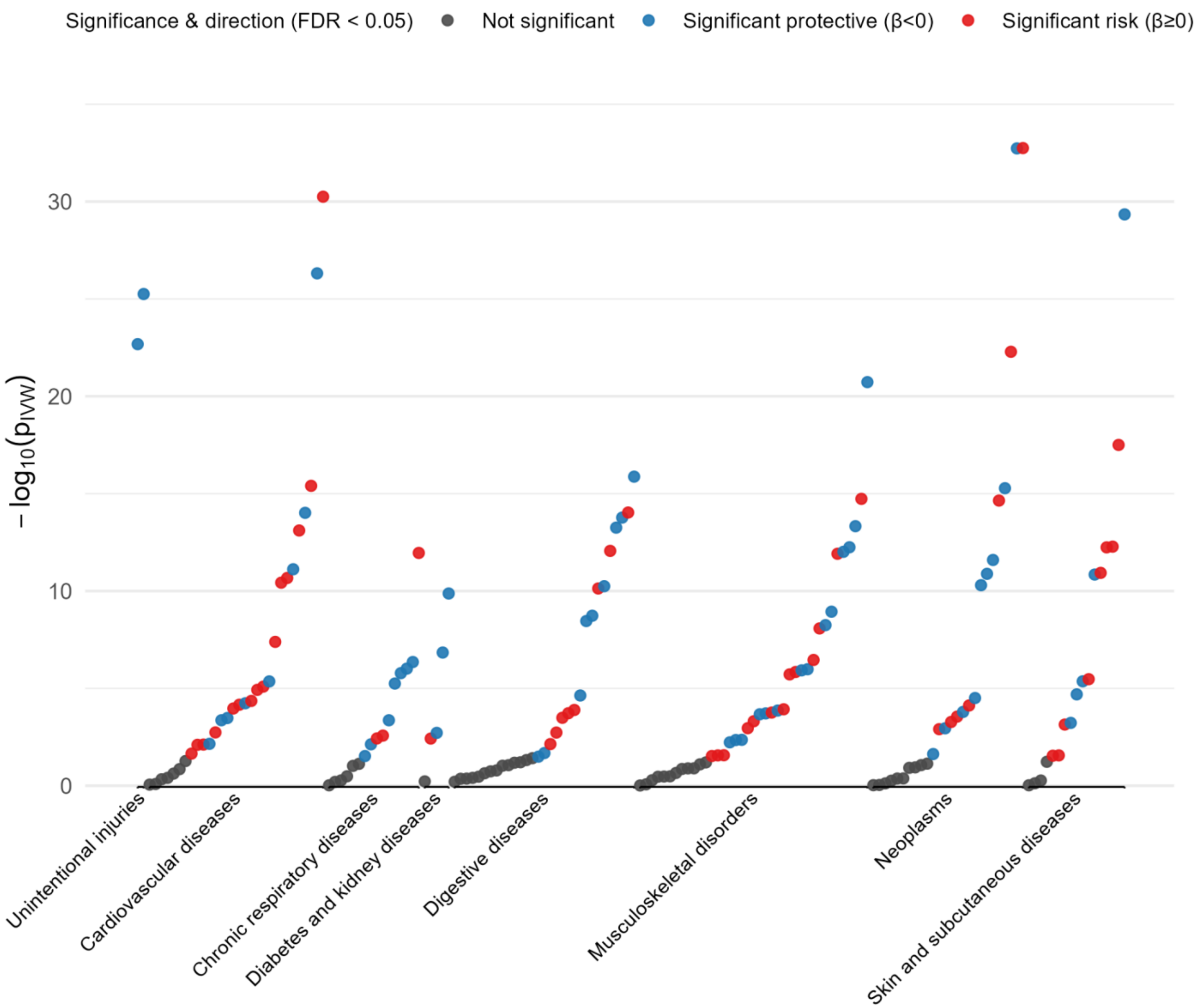


**Supplementary Figure 4.** Manhattan plot of GLP-1R expression effects on the phenome of age-related diseases in the female European population. Diseases are grouped by Global Burden of Disease category (level 2). Disease labels are applied to the lowest p-value effect in each category, and to significantly affected diseases in the most risk-affected and protective-affected categories. Red dots imply increased GLP-1R expression raises risk for the named disease outcome, blue dots imply increased GLP-1R expression lowers risk, and grey dots represent null findings. Statistical significance is defined as q < 0.05 after Benjamini-Hochberg false discovery rate adjustment. -log10(Pivw) represents the log-scaled p-value measuring the statistical significance of the measured effect.

Supplementary Figure 5. ARD-wide MR results for GLP1R, European male population.

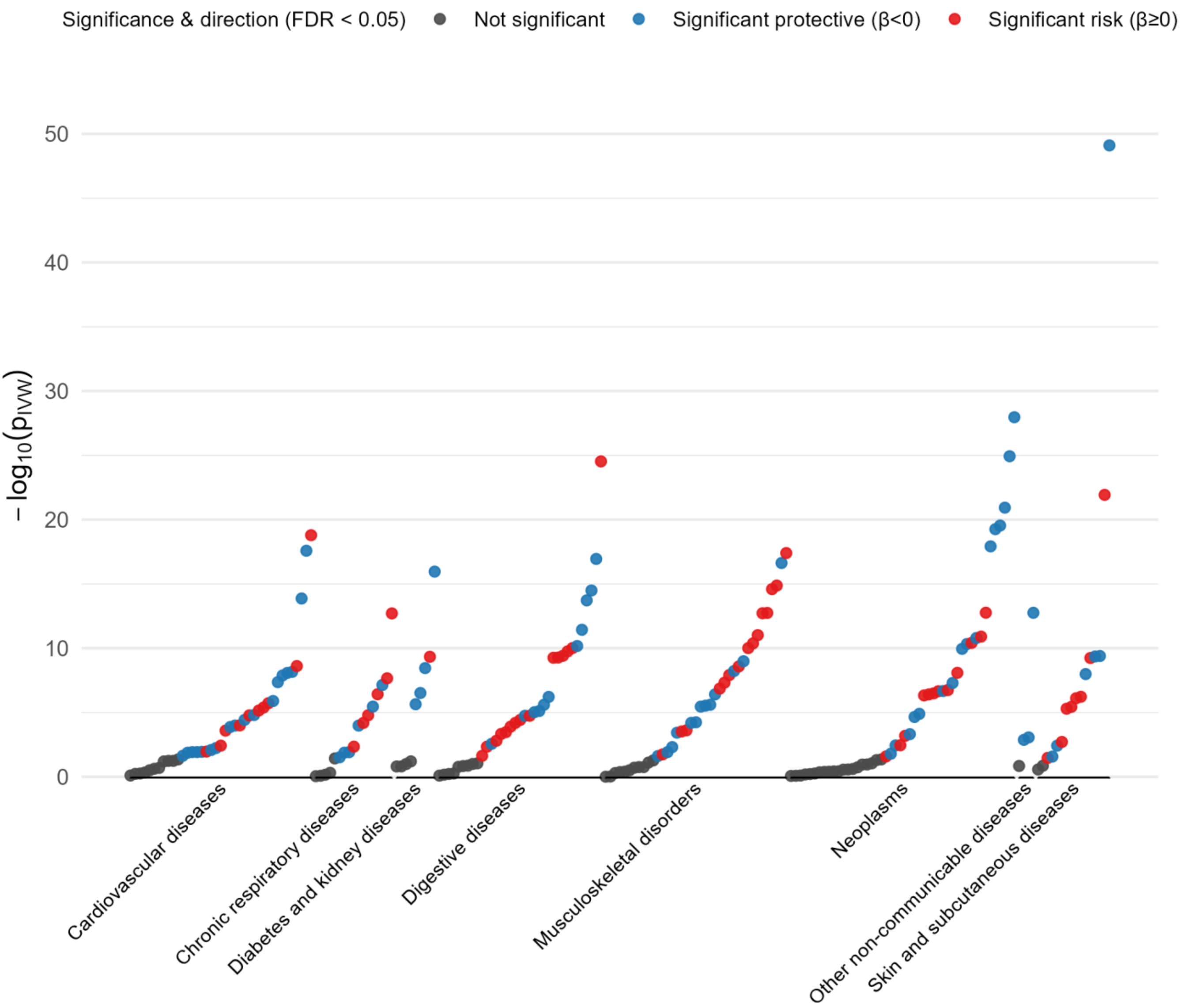


**Supplementary Figure 5.** Manhattan plot of GLP-1R expression effects on the phenome of age-related diseases in the male European population. Diseases are grouped by Global Burden of Disease category (level 2). Disease labels are applied to the lowest p-value effect in each category, and to significantly affected diseases in the most risk-affected and protective-affected categories. Red dots imply increased GLP-1R expression raises risk for the named disease outcome, blue dots imply increased GLP-1R expression lowers risk, and grey dots represent null findings. Statistical significance is defined as q < 0.05 after Benjamini-Hochberg false discovery rate adjustment. -log10(Pivw) represents the log-scaled p-value measuring the statistical significance of the measured effect.

Supplementary Figure 6. Permutation test for ageing-phenome wide effect of GLP1R, European both-sex population.

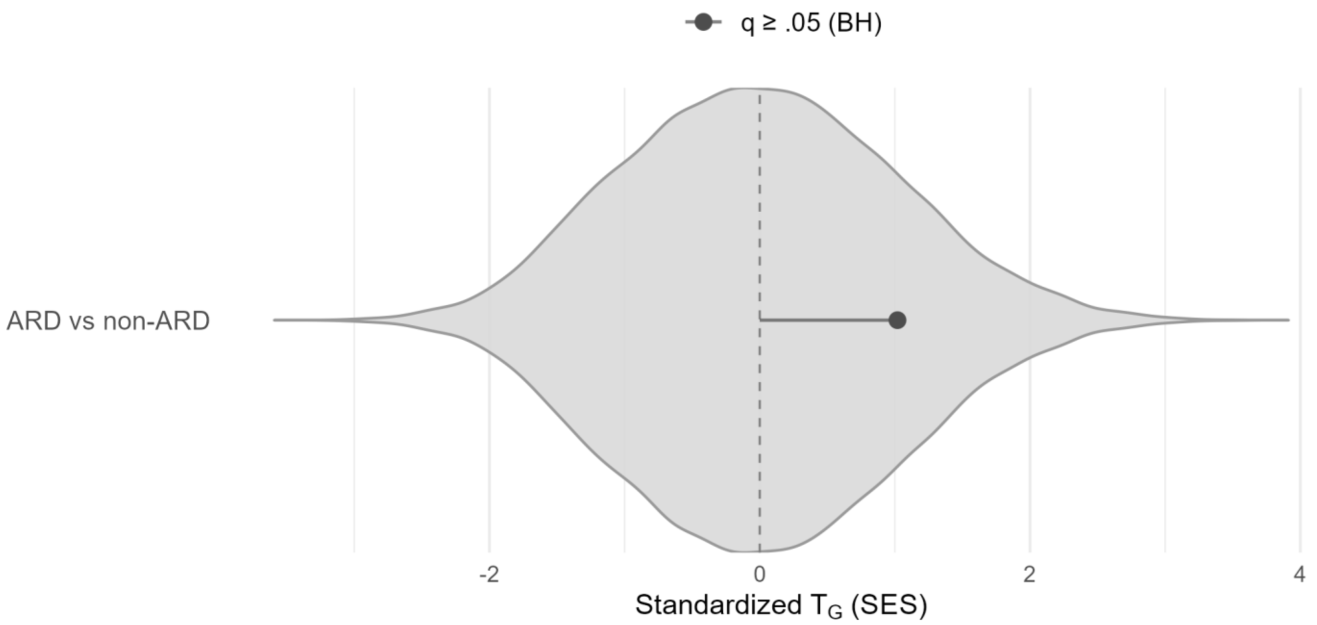


**Supplementary Figure 6.** Permutation testing for category-wide effects of GLP-1R expression on the both-sex European population. The single category contrasts age-related diseases (ARD) versus non-ARD outcomes. Red dots imply increased GLP-1R expression raises risk for the diseases in the named disease category, and grey dots represent null findings.

Supplementary Figure 7. Permutation test for effects of GLP1R on age-related disease, European female population.

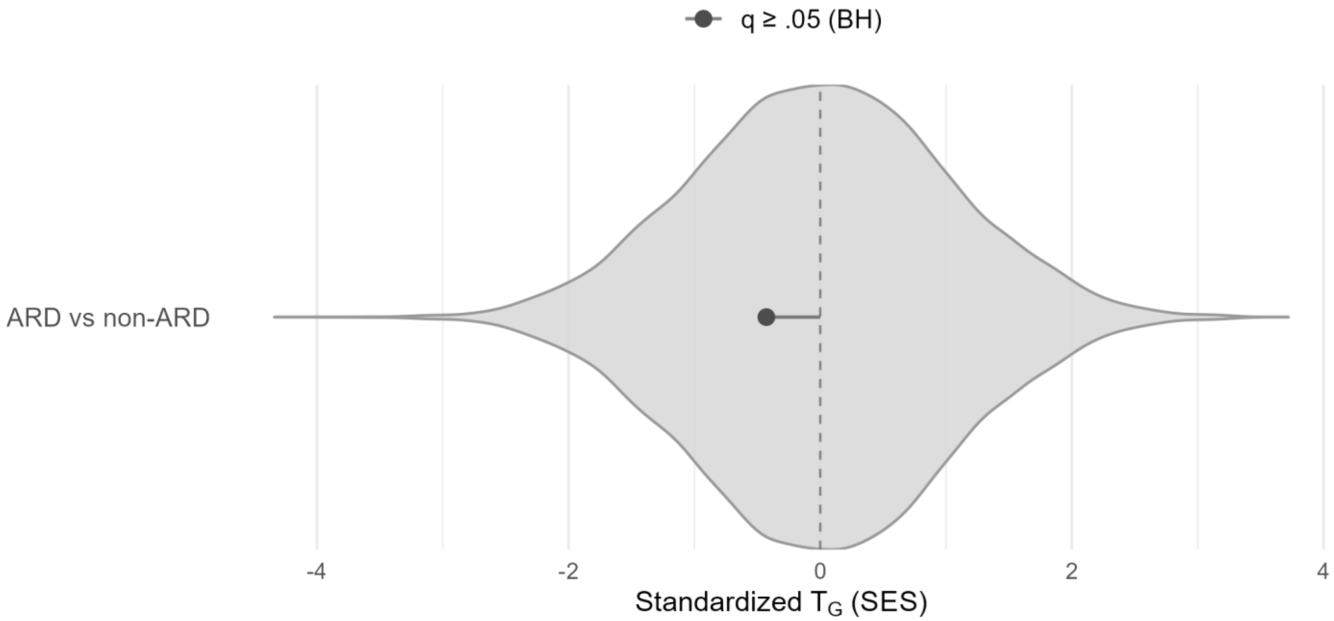


**Supplementary Figure 7.** Permutation testing for category-wide effects of GLP-1R expression on the female European population. The single category contrasts age-related diseases (ARD) versus non-ARD outcomes. Red dots imply increased GLP-1R expression raises risk for the diseases in the named disease category, and grey dots represent null findings. Statistical significance is defined as $q < 0.05$ after Benjamini-Hochberg false discovery rate adjustment.

Supplementary Figure 8. Permutation test for effects of GLP1R on age-related disease, European male population.

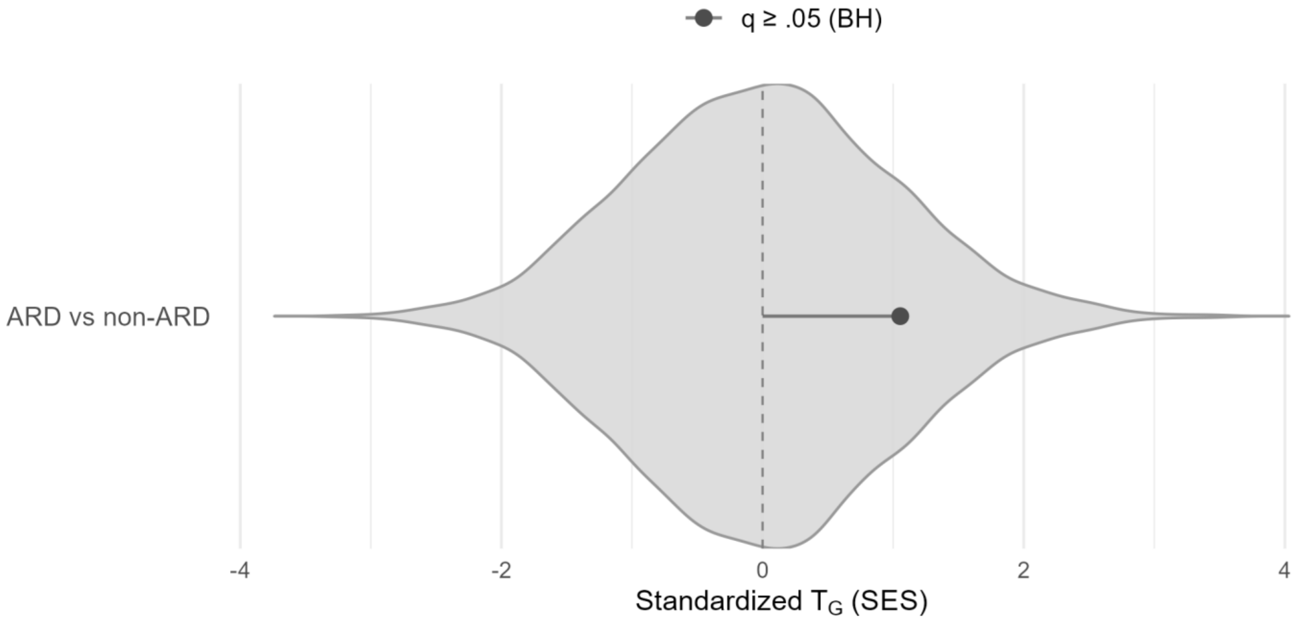


**Supplementary Figure 8.** Permutation testing for category-wide effects of GLP-1R expression on the male European population. The single category contrasts age-related diseases (ARD) versus non-ARD outcomes. Red dots imply increased GLP-1R expression raises risk for the diseases in the named disease category, and grey dots represent null findings. Statistical significance is defined as $q < 0.05$ after Benjamini-Hochberg false discovery rate adjustment.

Supplementary Figure 9. Permutation test for effects of GLP1R on level one GBD diseases, European both-sex population

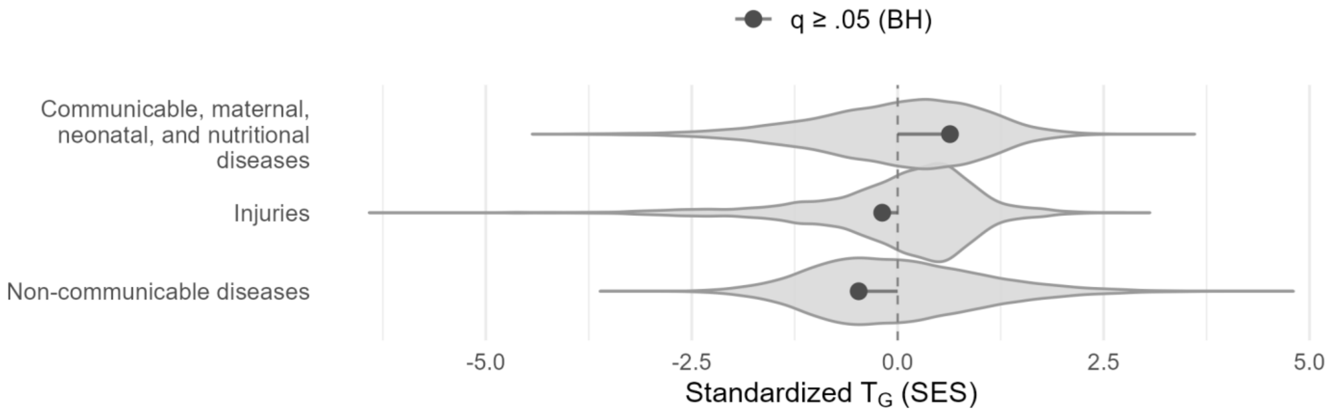


**Supplementary Figure 9.** Permutation testing for category-wide effects of GLP-1R expression on the both-sex European population. Categories are drawn from level 1 of the Global Burden of Disease clinical ontology. Red dots imply increased GLP-1R expression raises risk for the diseases in the named disease category, and grey dots represent null findings. Statistical significance is defined as $q < 0.05$ after Benjamini-Hochberg false discovery rate adjustment.

Supplementary Figure 10. Permutation test for effects of GLP1R on level two GBD diseases, European both sexes

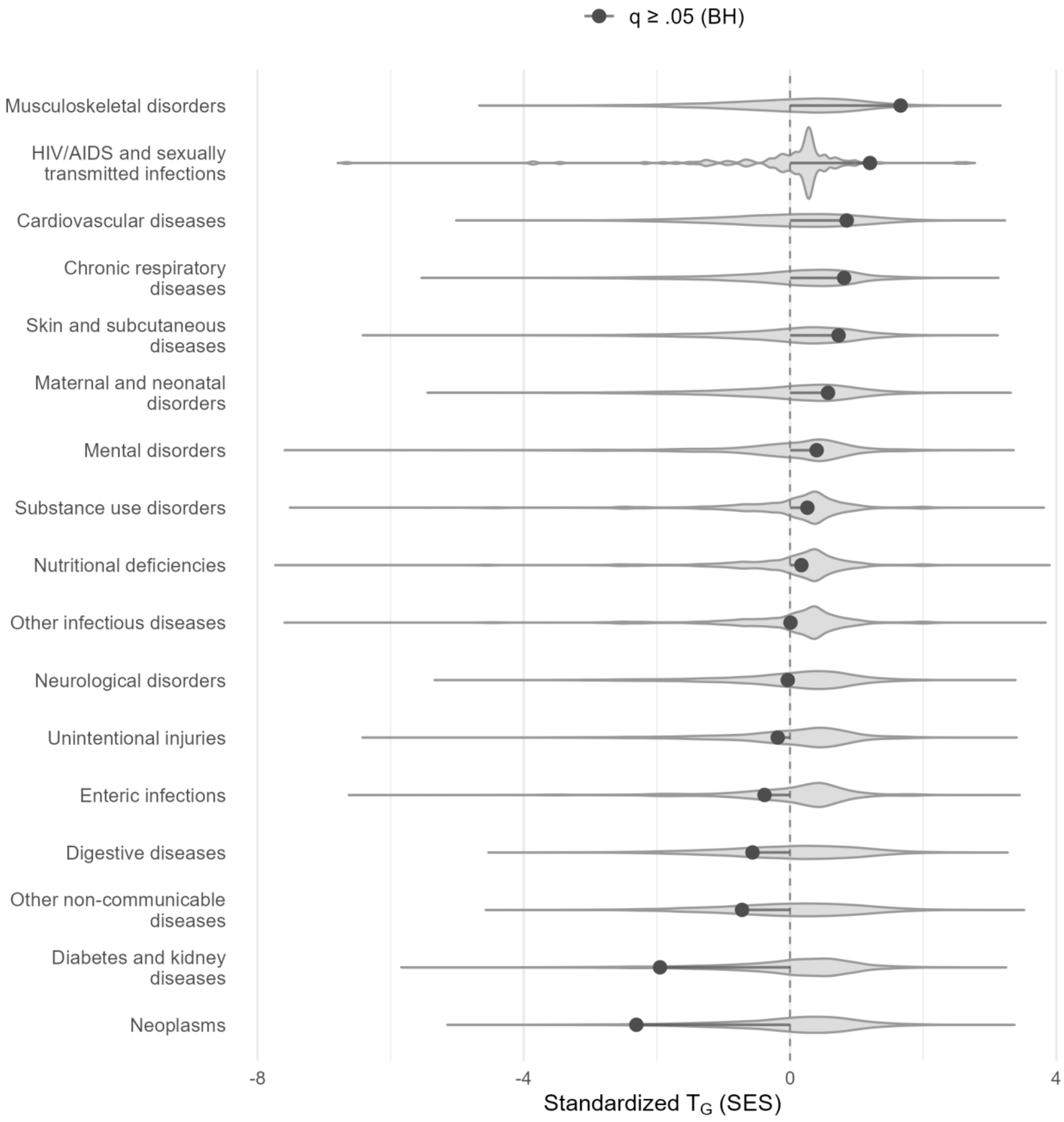


**Supplementary Figure 10.** Permutation testing for category-wide effects of GLP-1R expression on the both-sex European population. Categories are drawn from level 2 of the Global Burden of Disease clinical ontology. Red dots imply increased GLP-1R expression raises risk for the diseases in the named disease category, and grey dots represent null findings. Statistical significance is defined as q < 0.05 after Benjamini-Hochberg false discovery rate adjustment.

Supplementary Figure 11. Permutation test for effects of GLP1R on level three GBD diseases, European both-sex population.

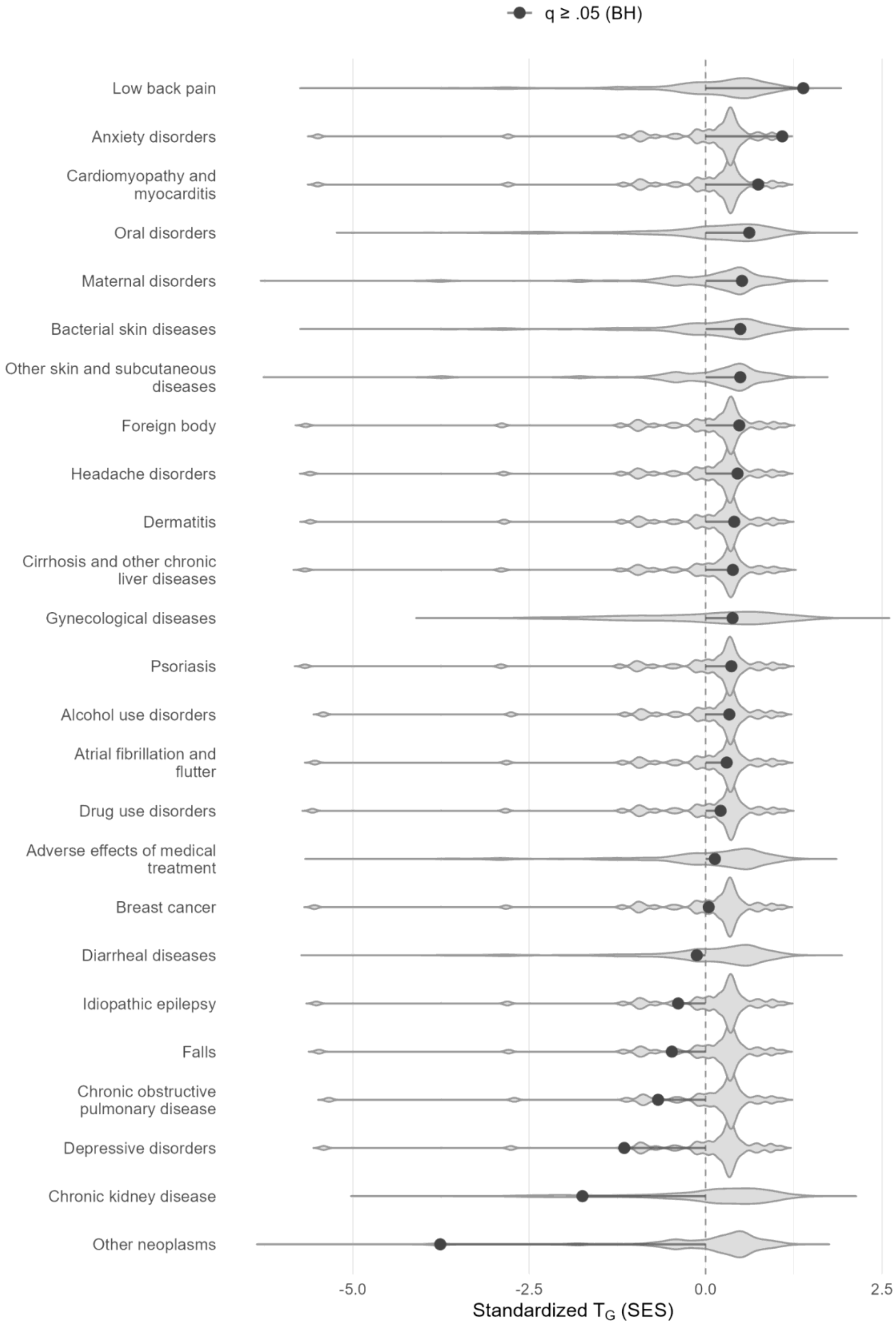


**Supplementary Figure 11.** Permutation testing for category-wide effects of GLP-1R expression on the both-sex European population. Categories are drawn from level 3 of the Global Burden of Disease clinical ontology. Grey dots represent null findings after Benjamini-Hochberg false discovery rate adjustment at $q < 0.05$.

Supplementary Figure 12. Permutation test for effects of GLP1R on level one GBD diseases, European female population.

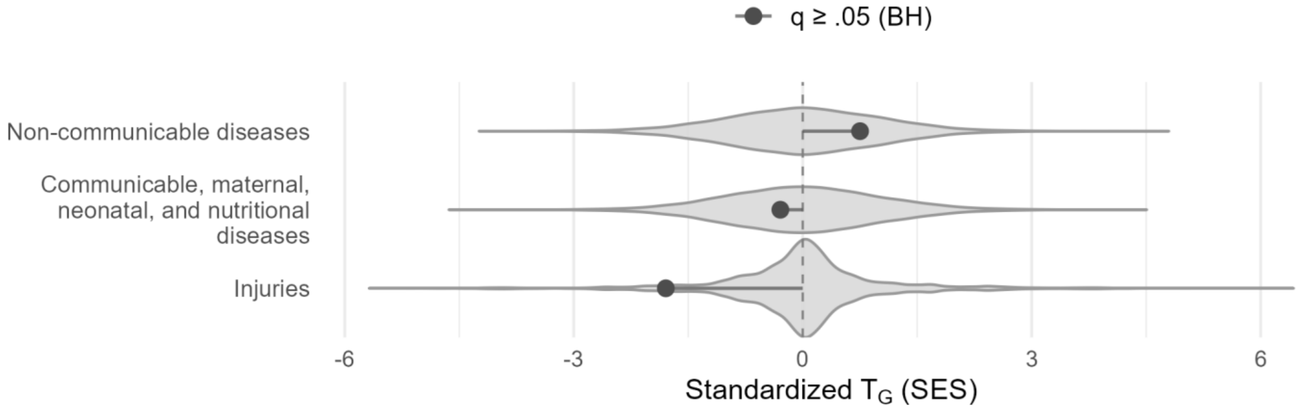


**Supplementary Figure 12.** Permutation testing for category-wide effects of GLP-1R expression on the female European population. Categories are drawn from level 1 of the Global Burden of Disease clinical ontology. Red dots imply increased GLP-1R expression raises risk for the diseases in the named disease category, and grey dots represent null findings. Statistical significance is defined as q < 0.05 after Benjamini-Hochberg false discovery rate adjustment.

Supplementary Figure 13. Permutation test for effects of GLP1R on level two GBD diseases, European female population.

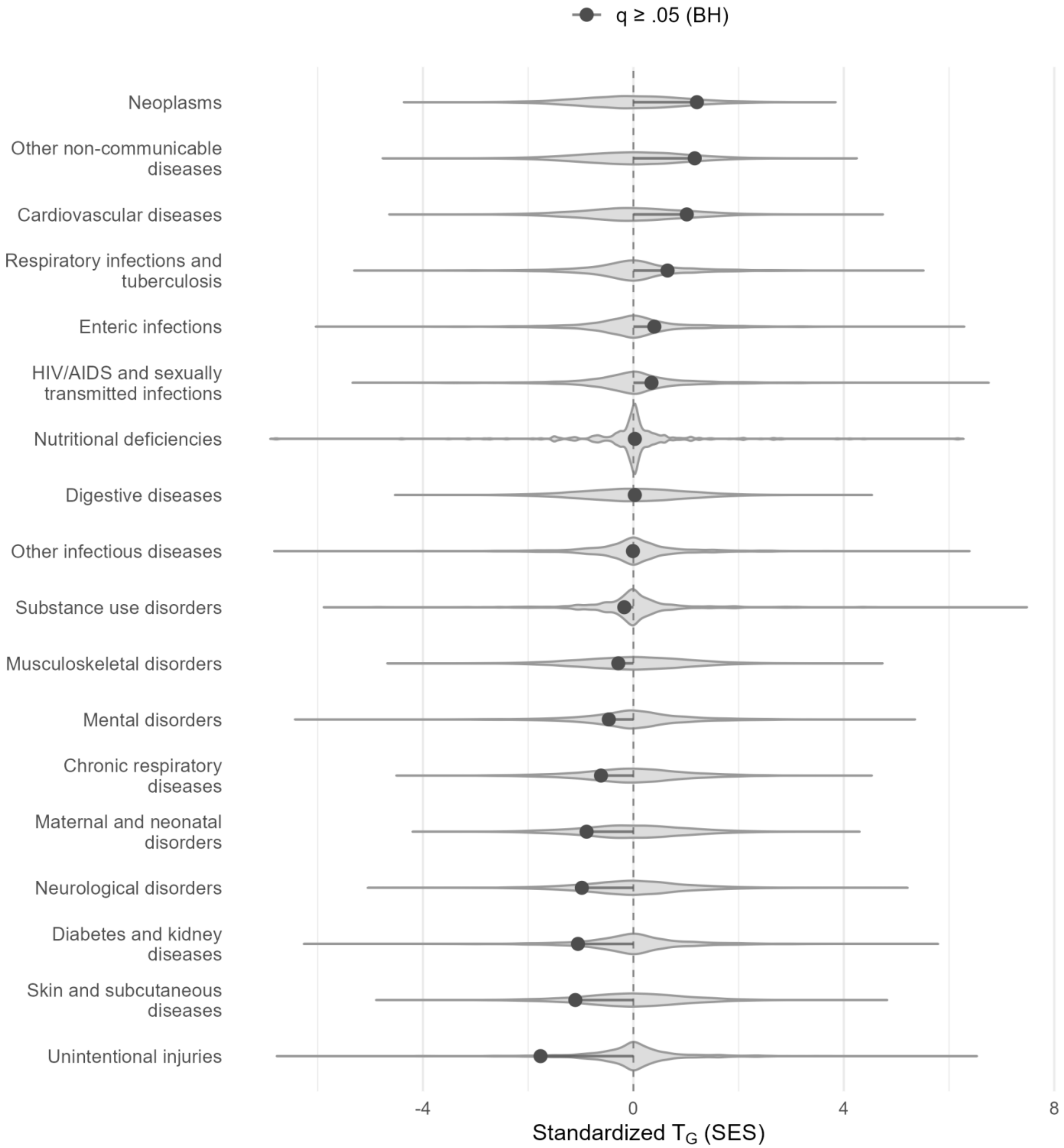


**Supplementary Figure 13.** Permutation testing for category-wide effects of GLP-1R expression on the female European population. Categories are drawn from level 2 of the Global Burden of Disease clinical ontology. Red dots imply increased GLP-1R expression raises risk for the diseases in the named disease category, and grey dots represent null findings. Statistical significance is defined as $q < 0.05$ after Benjamini-Hochberg false discovery rate adjustment.

Supplementary Figure 14. Permutation test for effects of GLP1R on level three GBD diseases, European female population.

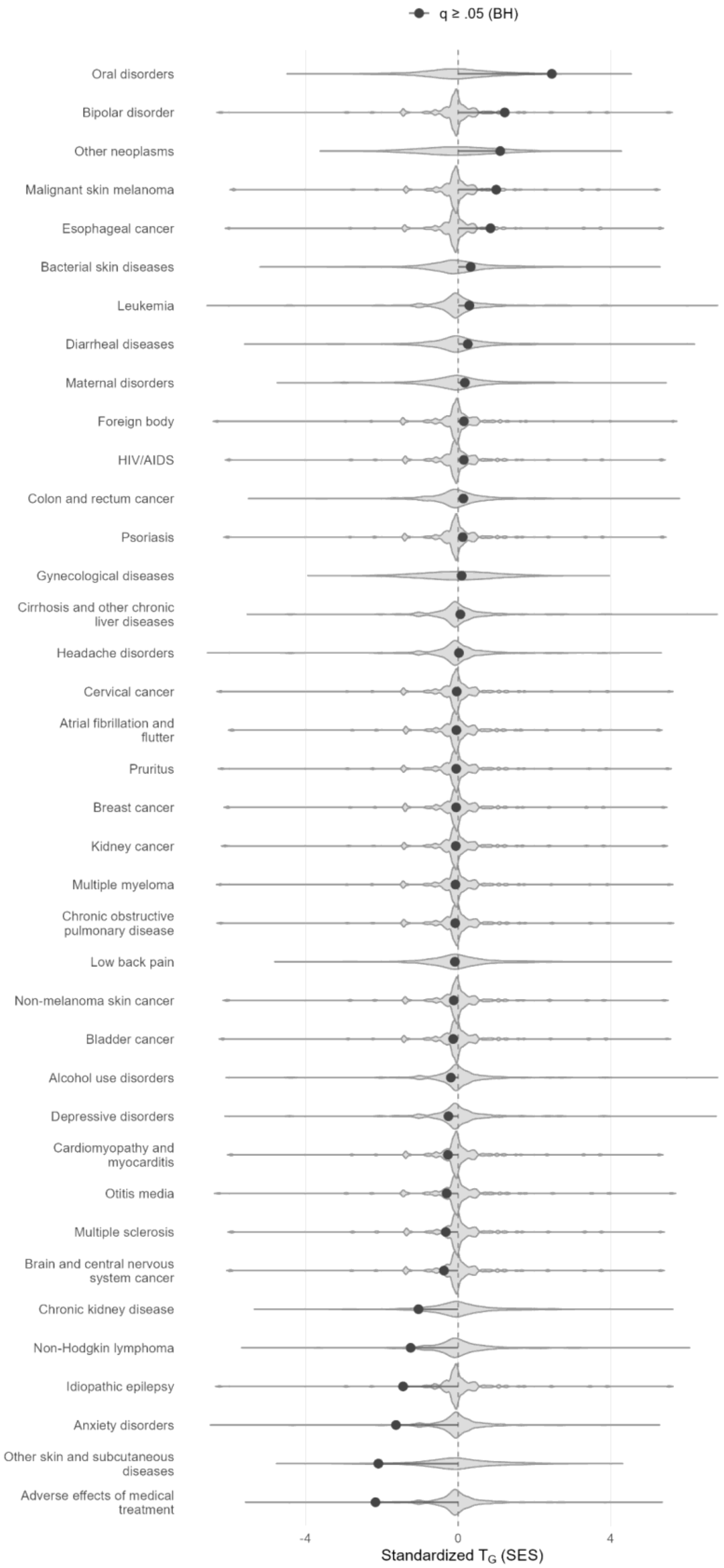


**Supplementary Figure 14.** Permutation testing for category-wide effects of GLP-1R expression on the female European population. Categories are drawn from level 3 of the Global Burden of Disease clinical ontology. Grey dots represent null findings after Benjamini-Hochberg false discovery rate adjustment at q < 0.05 .

Supplementary Figure 15. Permutation test for effects of GLP1R on level one GBD diseases, European male population.

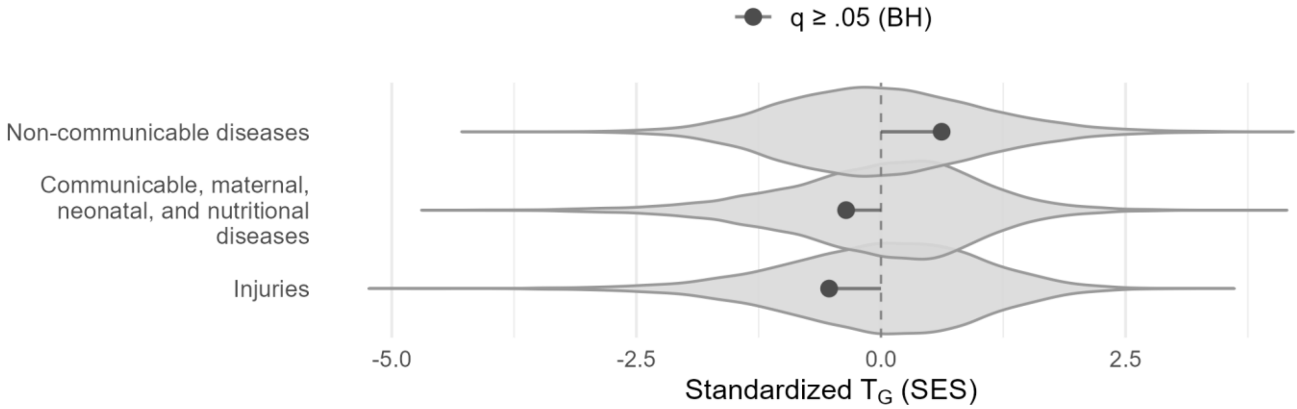


**Supplementary Figure 15.** Permutation testing for category-wide effects of GLP-1R expression on the male European population. Categories are drawn from level 1 of the Global Burden of Disease clinical ontology. Red dots imply increased GLP-1R expression raises risk for the diseases in the named disease category, and grey dots represent null findings. Statistical significance is defined as $q < 0.05$ after Benjamini-Hochberg false discovery rate adjustment.

Supplementary Figure 16. Permutation test for effects of GLP1R on level two GBD diseases, European male population.

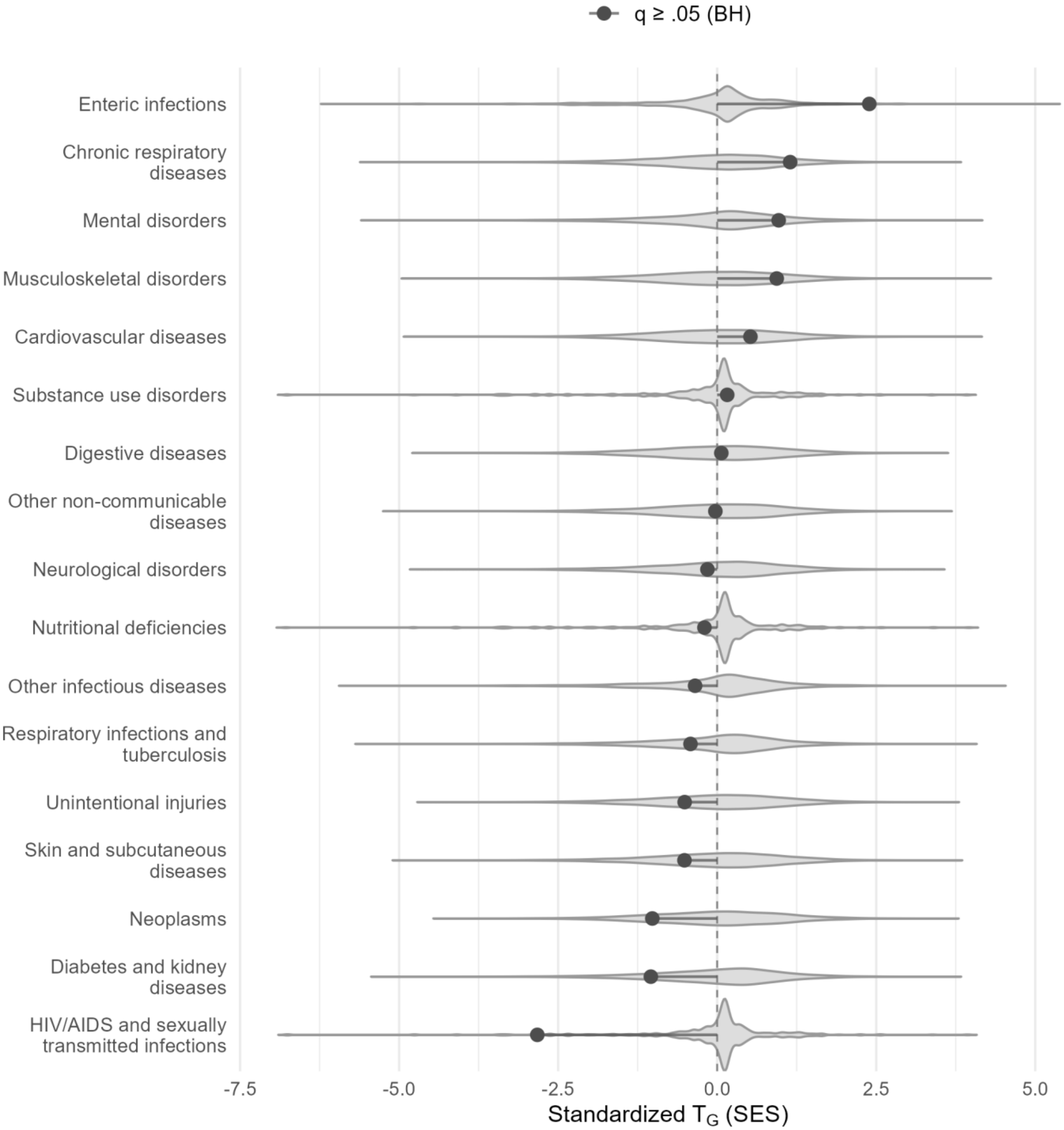


**Supplementary Figure 16.** Permutation testing for category-wide effects of GLP-1R expression on the male European population. Categories are drawn from level 2 of the Global Burden of Disease clinical ontology. Red dots imply increased GLP-1R expression raises risk for the diseases in the named disease category, and grey dots represent null findings. Statistical significance is defined as q < 0.05 after Benjamini-Hochberg false discovery rate adjustment.

Supplementary Figure 17. Permutation test for effects of GLP1R on level three GBD diseases, European male population.

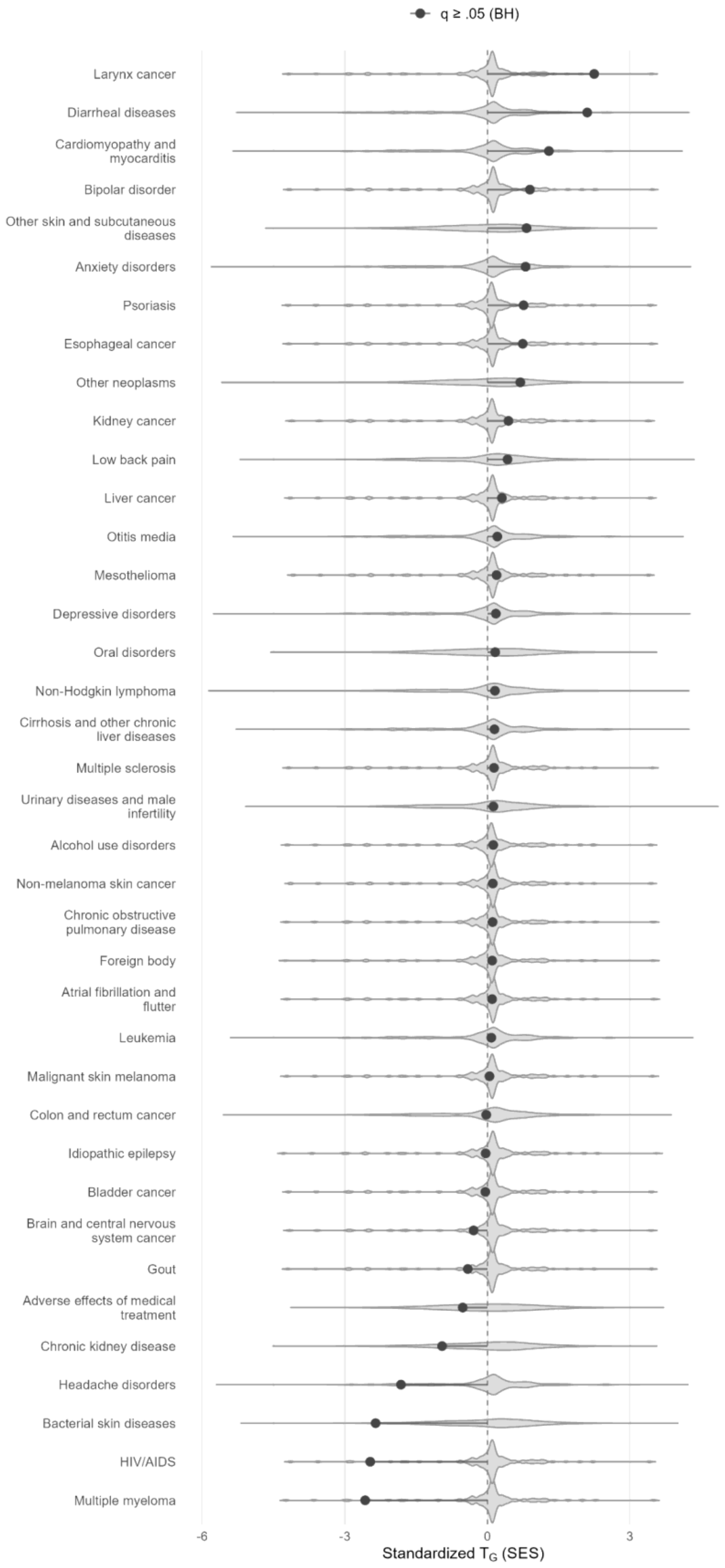


**Supplementary Figure 17.** Permutation testing for category-wide effects of GLP-1R expression on the male European population. Categories are drawn from level 3 of the Global Burden of Disease clinical ontology. Grey dots represent null findings after Benjamini-Hochberg false discovery rate adjustment (q < 0.05).

Supplementary Figure 18. Phenome-wide MR results for CETP, European female population.

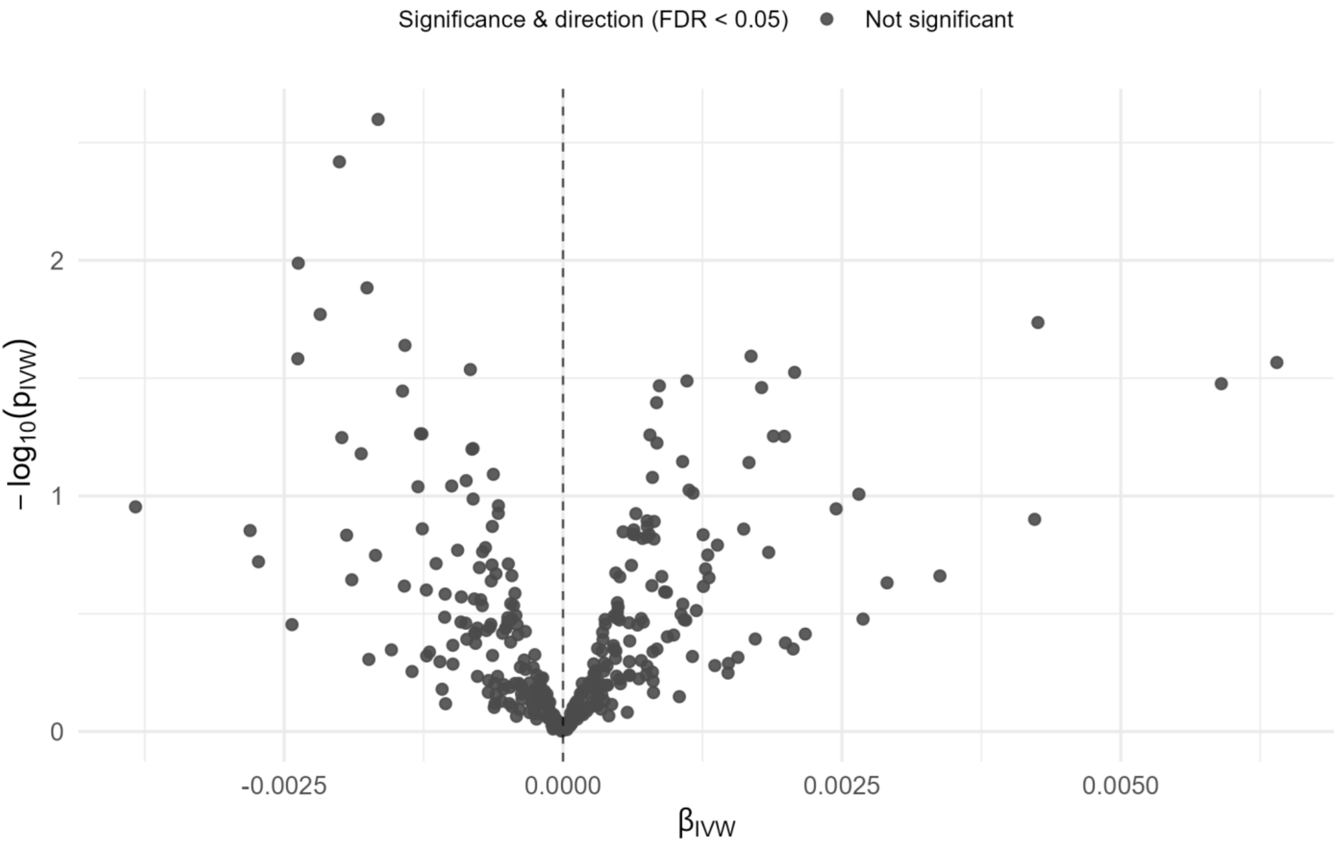


**Supplementary Figure 18.** Volcano plot results of phenome-wide Mendelian randomization of CETP concentration in female European population. "MR" refers to Mendelian Randomization testing. "CETP concentration" refers to genetically proxied CETP concentration. "FDR" refers to Benjamini-Hochberg false discovery rate correction. "βivw" refers to the causal effect of one standard deviation increase of CETP concentration on log-odds ratio of disease diagnosis as estimated using the inverse variance weighted method. The dotted line refers to a log-odds ratio of zero, implying no effect change in disease risk resulting from increased CETP concentration. Disease labels are applied to selected novel Mendelian randomization findings with selective labelling. Red dots imply increased CETP concentration raises risk for the named disease outcome, blue dots imply increased CETP concentration lowers risk, and grey dots represent null findings. Statistical significance is defined as q < 0.05 after Benjamini-Hochberg false discovery rate adjustment.

Supplementary Figure 19. Phenome-wide MR results for CETP, European male population.

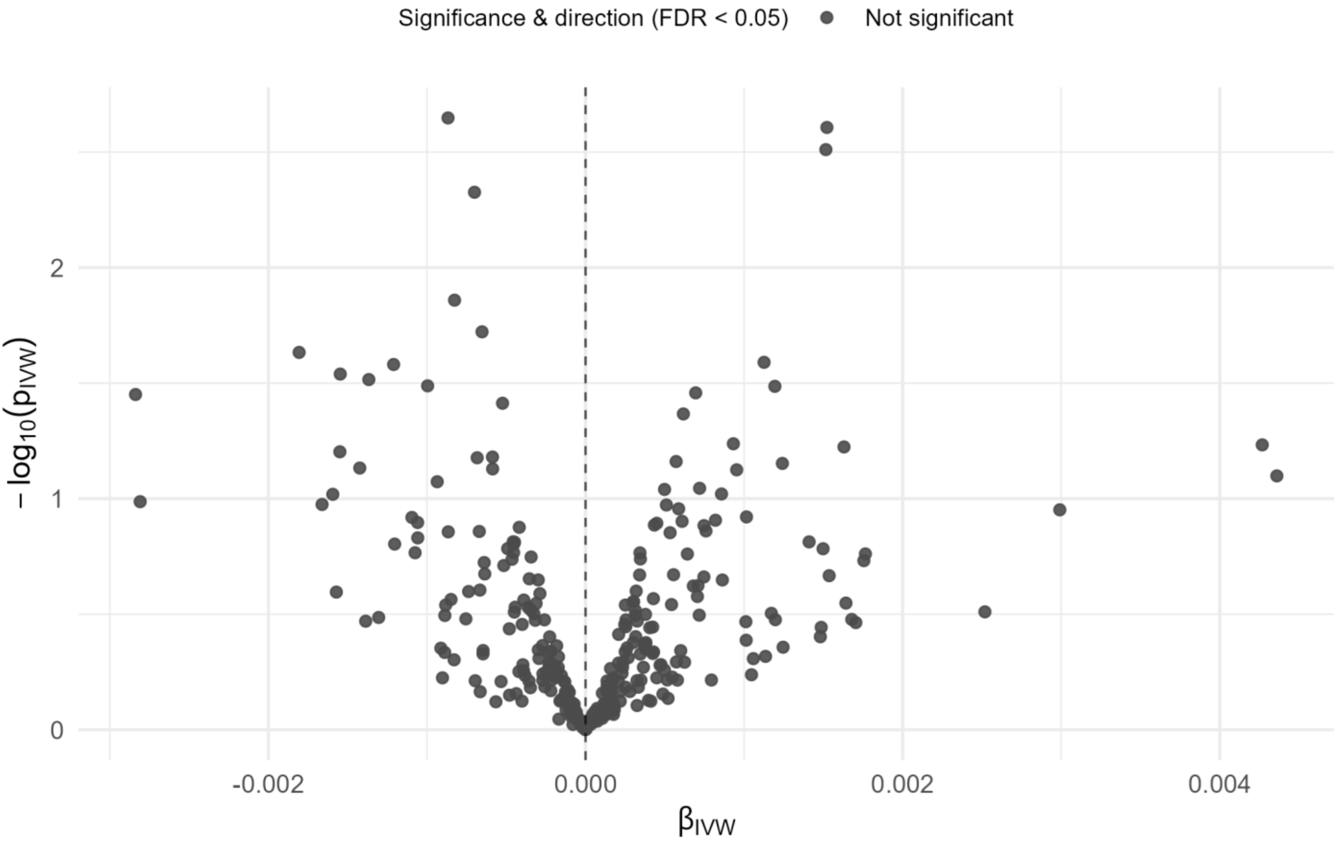


**Supplementary Figure 19.** Volcano plot results of phenome-wide Mendelian randomization of CETP concentration in male European population. "MR" refers to Mendelian Randomization testing. "CETP concentration" refers to genetically proxied CETP concentration. "FDR" refers to Benjamini-Hochberg false discovery rate correction. "βivw" refers to the causal effect of one standard deviation increase of CETP concentration on log-odds ratio of disease diagnosis as estimated using the inverse variance weighted method. The dotted line refers to a log-odds ratio of zero, implying no effect change in disease risk resulting from increased CETP concentration. Disease labels are applied to selected novel Mendelian randomization findings with selective labelling. Red dots imply increased CETP concentration raises risk for the named disease outcome, blue dots imply increased CETP concentration lowers risk, and grey dots represent null findings. Statistical significance is defined as $q < 0.05$ after Benjamini-Hochberg false discovery rate adjustment.

Supplementary Figure 20. Permutation test for effects of CETP on age-related disease, European both-sex population.

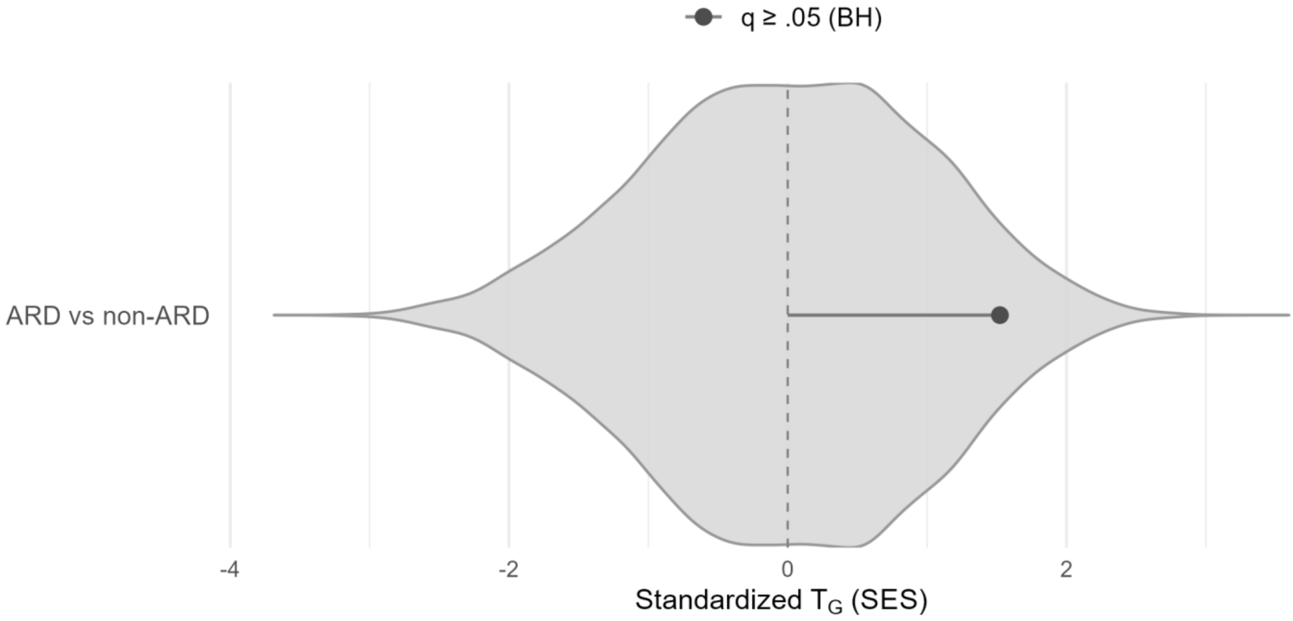


**Supplementary Figure 20.** Permutation testing for category-wide effects of CETP concentration on the both-sex European population. The single category contrasts age-related diseases (ARD) versus non-ARD outcomes. Red dots imply increased CETP concentration raises risk for the diseases in the named disease category, and grey dots represent null findings. Statistical significance is defined as q < 0.05 after Benjamini-Hochberg false discovery rate adjustment.

Supplementary Figure 21. Permutation test for effects of CETP on level one GBD diseases, both-sex population.

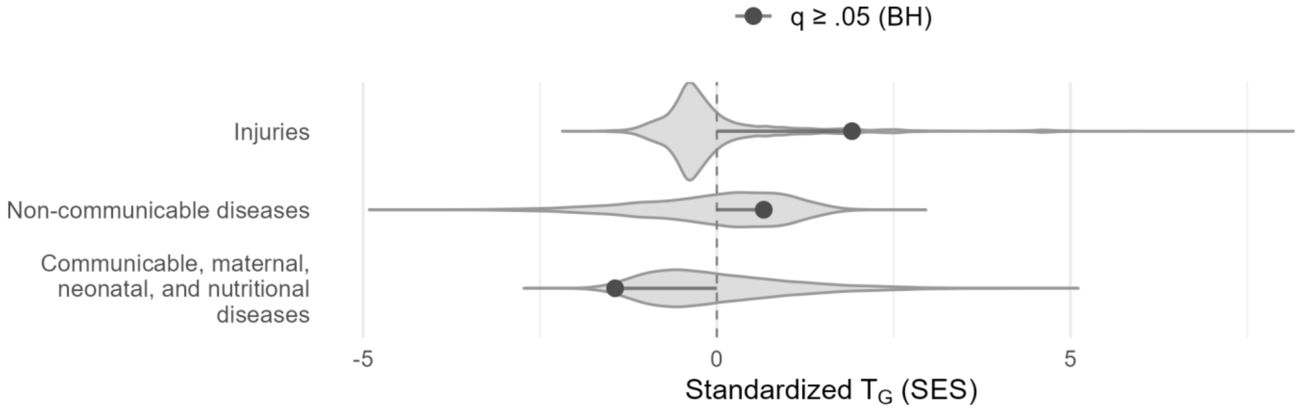


**Supplementary Figure 21.** Permutation testing for category-wide effects of CETP concentration on the both-sex population. Categories are drawn from level 1 of the Global Burden of Disease clinical ontology. Red dots imply increased CETP concentration raises risk for the diseases in the named disease category, and grey dots represent null findings. Statistical significance is defined as $q < 0.05$ after Benjamini-Hochberg false discovery rate adjustment.

Supplementary Figure 22. Permutation test for effects of CETP on level three GBD diseases, European both-sex population.

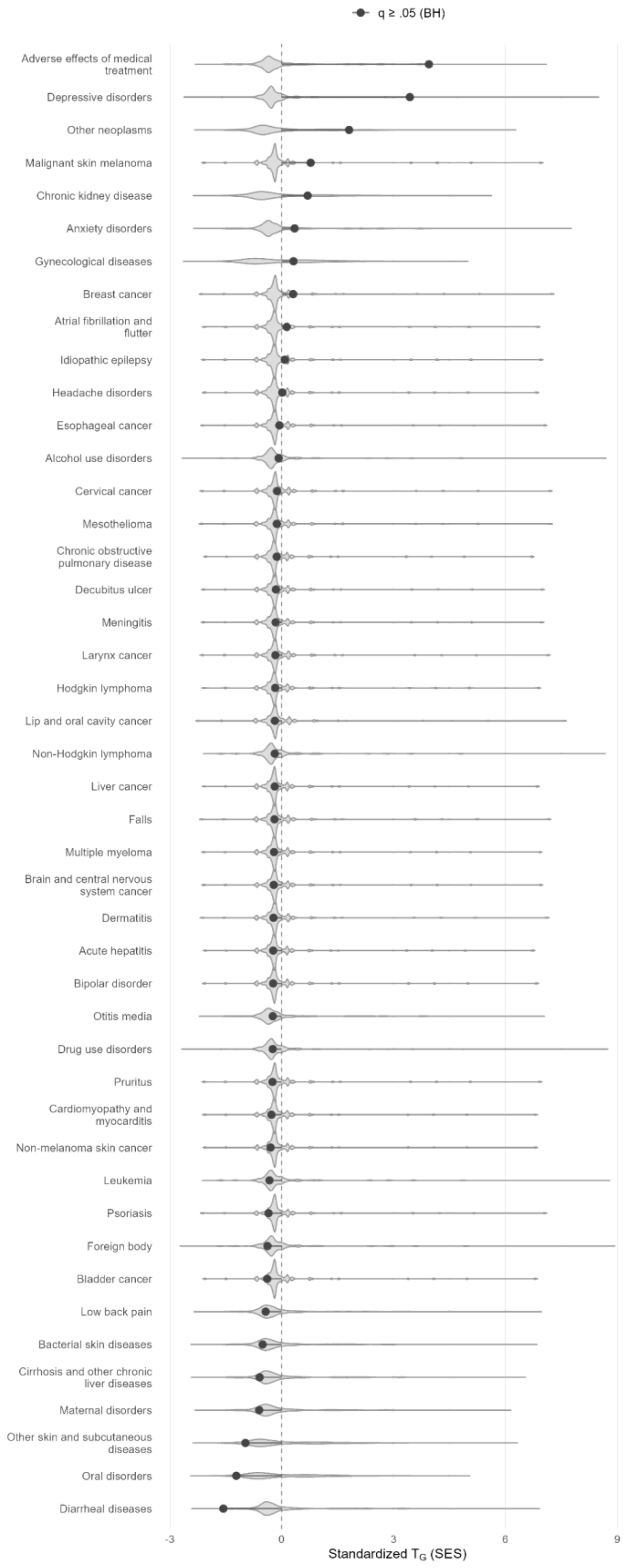


**Supplementary Figure 22.** Permutation testing for category-wide effects of CETP concentration on the both-sex European population. Categories are drawn from level 3 of the Global Burden of Disease clinical ontology. Grey dots represent null findings after Benjamini-Hochberg false discovery rate adjustment (q < 0.05).

Supplementary Figure 23. Phenome-wide MR results for ambidextrousness, European both-sex population.

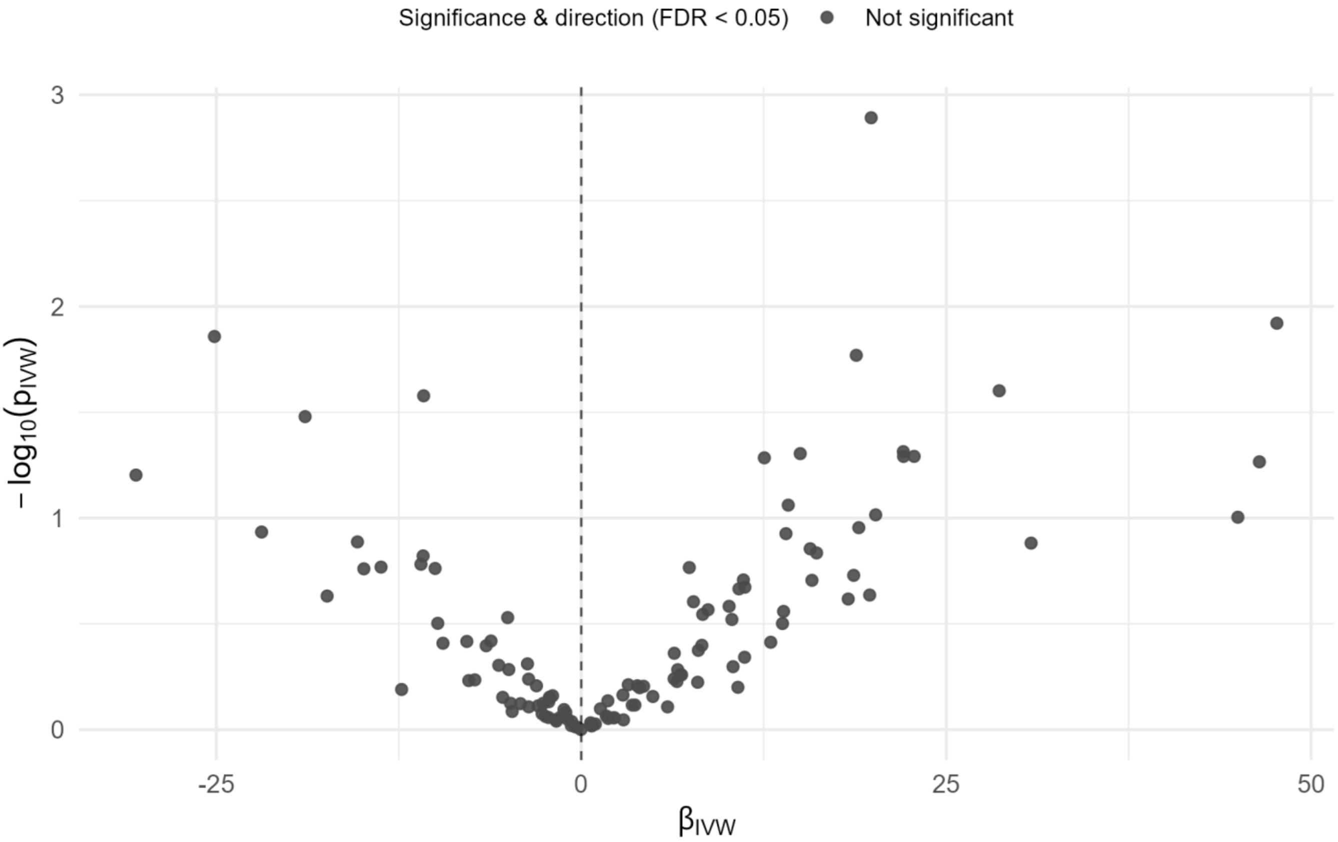


**Supplementary Figure 23.** Volcano plot results of phenome-wide Mendelian randomization of ambidextrousness in both-sex European population. “MR” refers to Mendelian Randomization testing. “ambidextrousness” refers to genetically proxied liability to ambidextrousness. “FDR” refers to Benjamini-Hochberg false discovery rate correction. “βivw” refers to the causal effect of one standard deviation increase of ambidextrousness on log-odds ratio of disease diagnosis as estimated using the inverse variance weighted method. The dotted line refers to a log-odds ratio of zero, implying no effect change in disease risk resulting from increased ambidextrousness. Disease labels are applied to selected novel Mendelian randomization findings with selective labelling. Red dots imply increased ambidextrousness raises risk for the named disease outcome, blue dots imply increased ambidextrousness lowers risk, and grey dots represent null findings. Statistical significance is defined as $q < 0.05$ after Benjamini-Hochberg false discovery rate adjustment.

Supplementary Figure 24. MR-PheWAS results for smoking initiation in both-sex Europeans (no pleiotropy filtering).

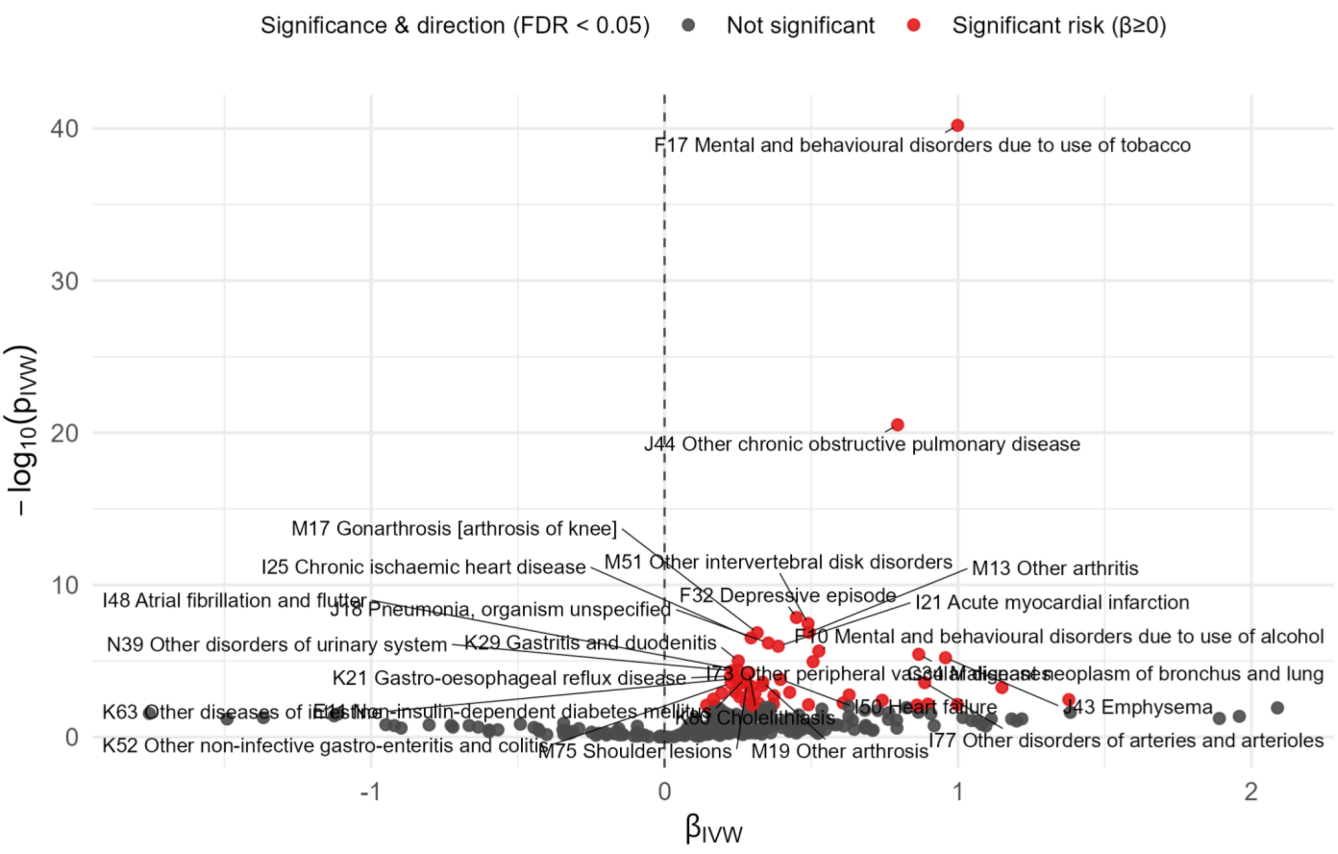


**Supplementary Figure 24.** Volcano plot results of phenome-wide Mendelian randomization of smoking initiation in both-sex European population. “MR” refers to Mendelian Randomization testing. “smoking initiation” refers to genetically proxied smoking initiation. “FDR” refers to Benjamini-Hochberg false discovery rate correction. “βivw” refers to the causal effect of one standard deviation increase of smoking initiation on log-odds ratio of disease diagnosis as estimated using the inverse variance weighted method. The dotted line refers to a log-odds ratio of zero, implying no effect change in disease risk resulting from increased smoking initiation. Disease labels are applied to selected novel Mendelian randomization findings with selective labelling. Red dots imply increased smoking initiation raises risk for the named disease outcome, blue dots imply increased smoking initiation lowers risk, and grey dots represent null findings. Statistical significance is defined as q < 0.05 after Benjamini-Hochberg false discovery rate adjustment.

Supplementary Figure 25. ARD-wide MR results for smoking initiation in both-sex Europeans (no pleiotropy filtering).

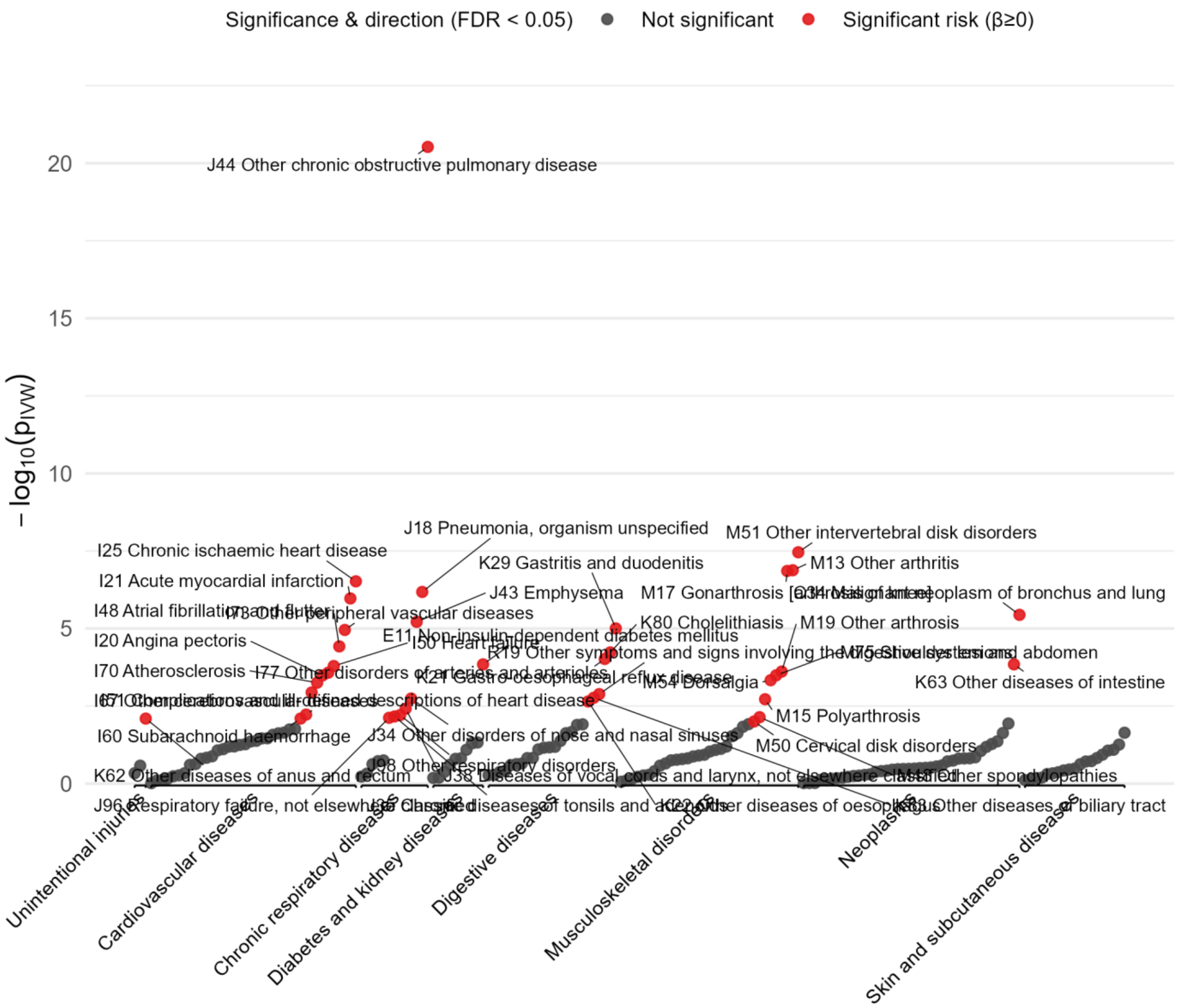


**Supplementary Figure 25.** Manhattan plot of smoking initiation effects on the phenome of age-related diseases in the both-sex European population. Diseases are grouped by Global Burden of Disease category (level 2). Disease labels are applied to the lowest p-value effect in each category, and to significantly affected diseases in the most risk-affected and protective-affected categories. Red dots imply increased smoking initiation raises risk for the named disease outcome, blue dots imply increased smoking initiation lowers risk, and grey dots represent null findings. Statistical significance is defined as q < 0.05 after Benjamini-Hochberg false discovery rate adjustment. -log10(Pivw) represents the log-scaled p-value measuring the statistical significance of the measured effect.

Supplementary Figure 26. Permutation tests for ageing phenome-wide effect of smoking initiation in both-sex Europeans (no pleiotropy filtering).

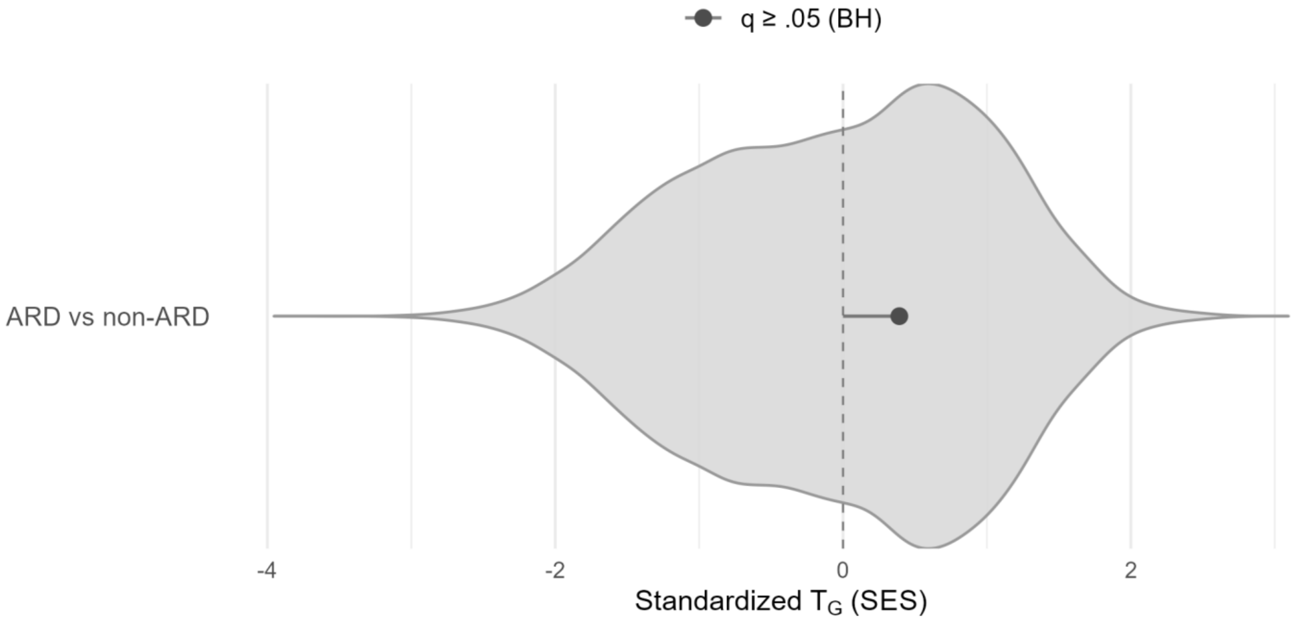


**Supplementary Figure 26.** Permutation testing for category-wide effects of smoking initiation on the both-sex European population. The single category contrasts age-related diseases (ARD) versus non-ARD outcomes. Red dots imply increased smoking initiation raises risk for the diseases in the named disease category, and grey dots represent null findings.

Supplementary Figure 27. Permutation test for effects of smoking initiation on level one GBD diseases, European both-sex population (no pleiotropy filtering).

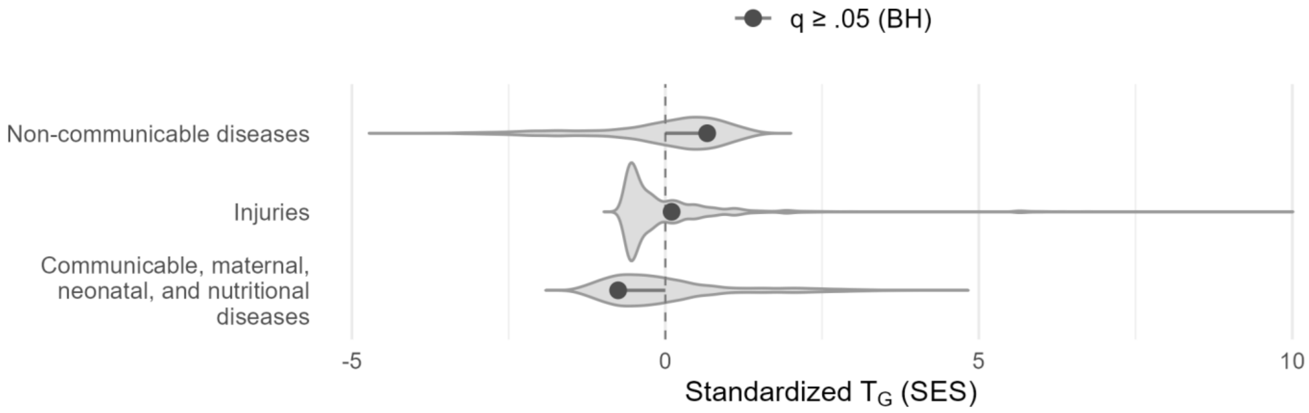


**Supplementary Figure 27.** Permutation testing for category-wide effects of smoking initiation on the both-sex European population. Categories are drawn from level 1 of the Global Burden of Disease clinical ontology. Red dots imply increased smoking initiation raises risk for the diseases in the named disease category, and grey dots represent null findings. Statistical significance is defined as q < 0.05 after Benjamini-Hochberg false discovery rate adjustment.

Supplementary Figure 28. Permutation test for effects of smoking initiation on level two GBD diseases, European both-sex population (no pleiotropy filtering).

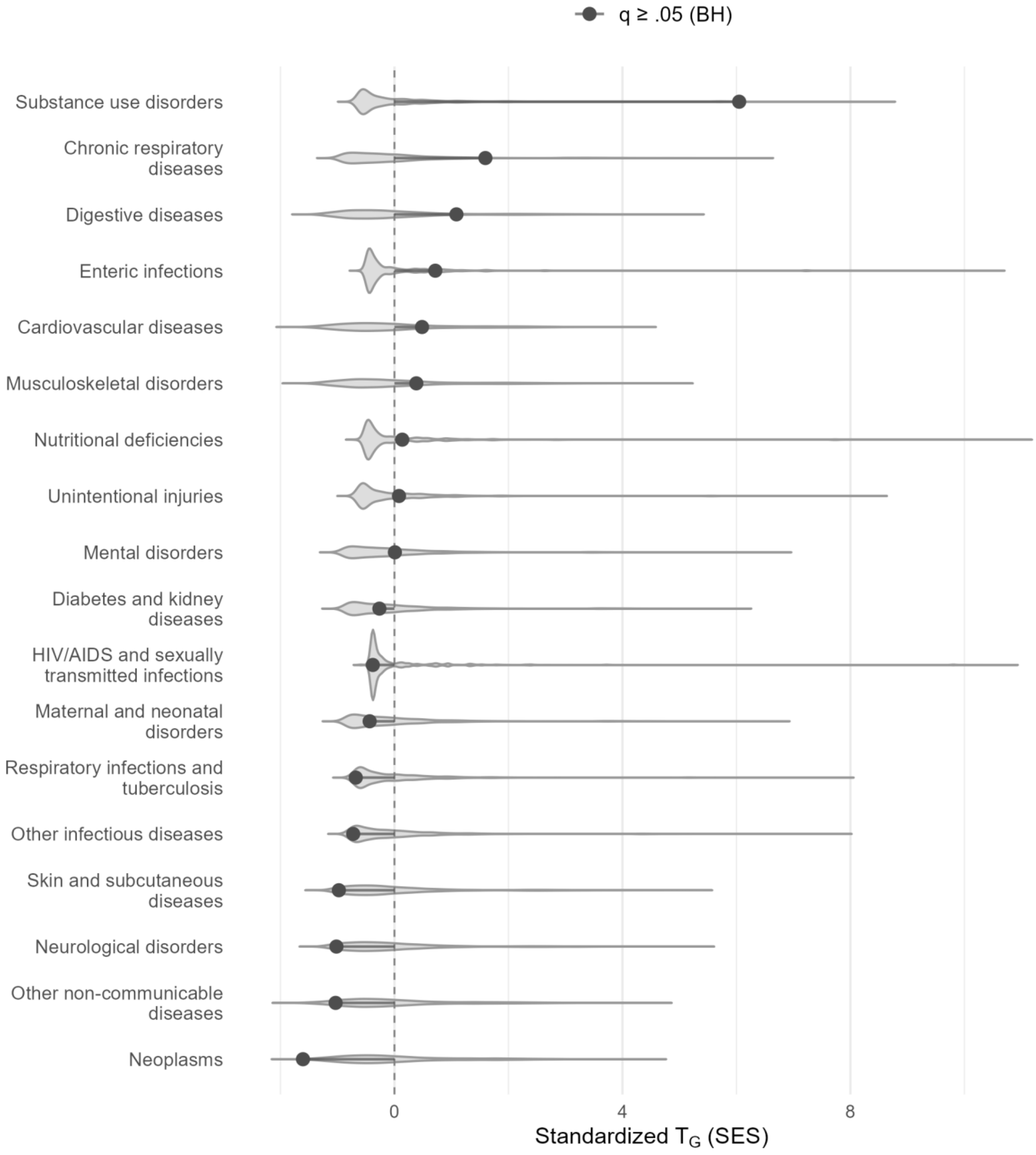


**Supplementary Figure 28.** Permutation testing for category-wide effects of smoking initiation on the both-sex European population. Categories are drawn from level 2 of the Global Burden of Disease clinical ontology. Red dots imply increased smoking initiation raises risk for the diseases in the named disease category, and grey dots represent null findings. Statistical significance is defined as q < 0.05 after Benjamini-Hochberg false discovery rate adjustment.

Supplementary Figure 29. Permutation test for effects of smoking initiation on level three GBD diseases, European both-sex population (no pleiotropy filtering).

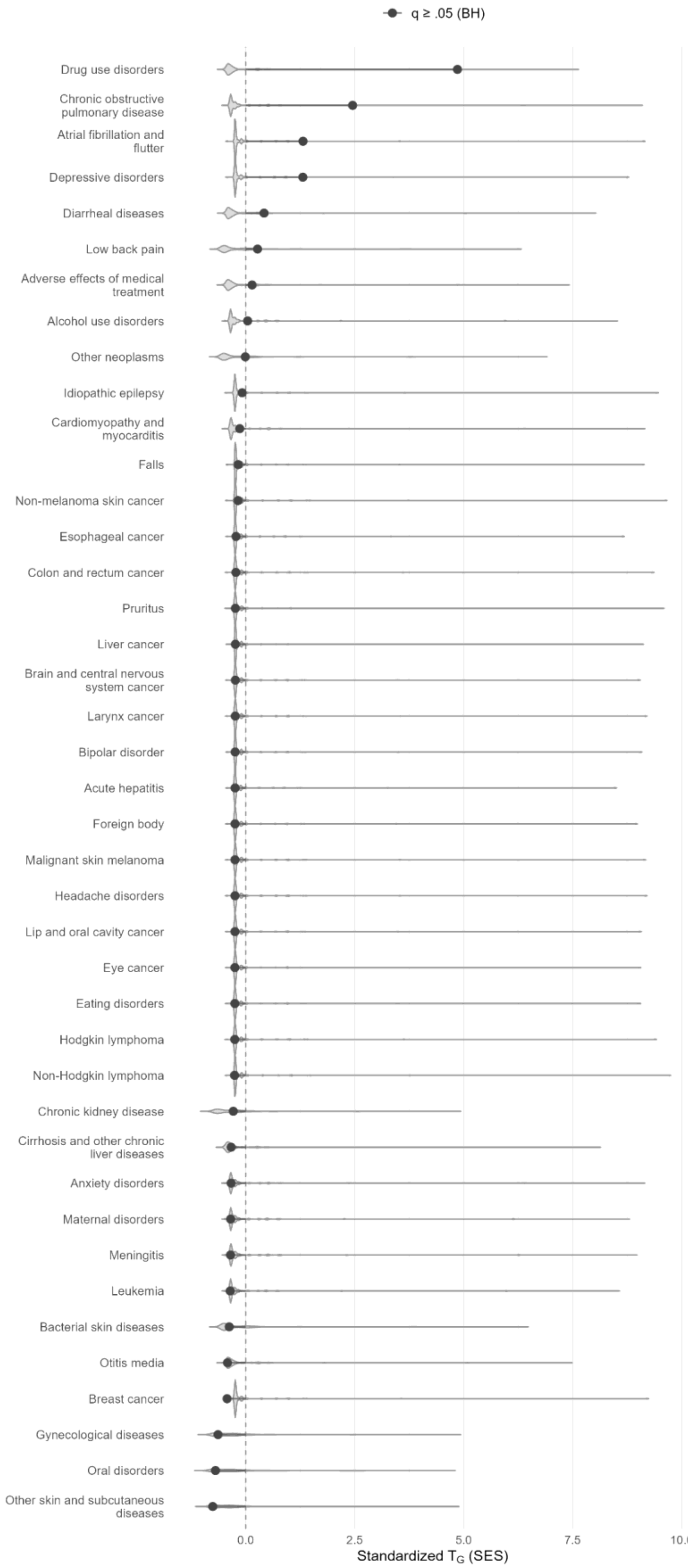


**Supplementary Figure 29.** Permutation testing for category-wide effects of smoking initiation on the both-sex European population. Categories are drawn from level 3 of the Global Burden of Disease clinical ontology. Grey dots represent null findings after Benjamini-Hochberg false discovery rate adjustment (q < 0.05).

Supplementary Figure 30. Phenome-wide MR results for smoking initiation, East Asian both-sex population.

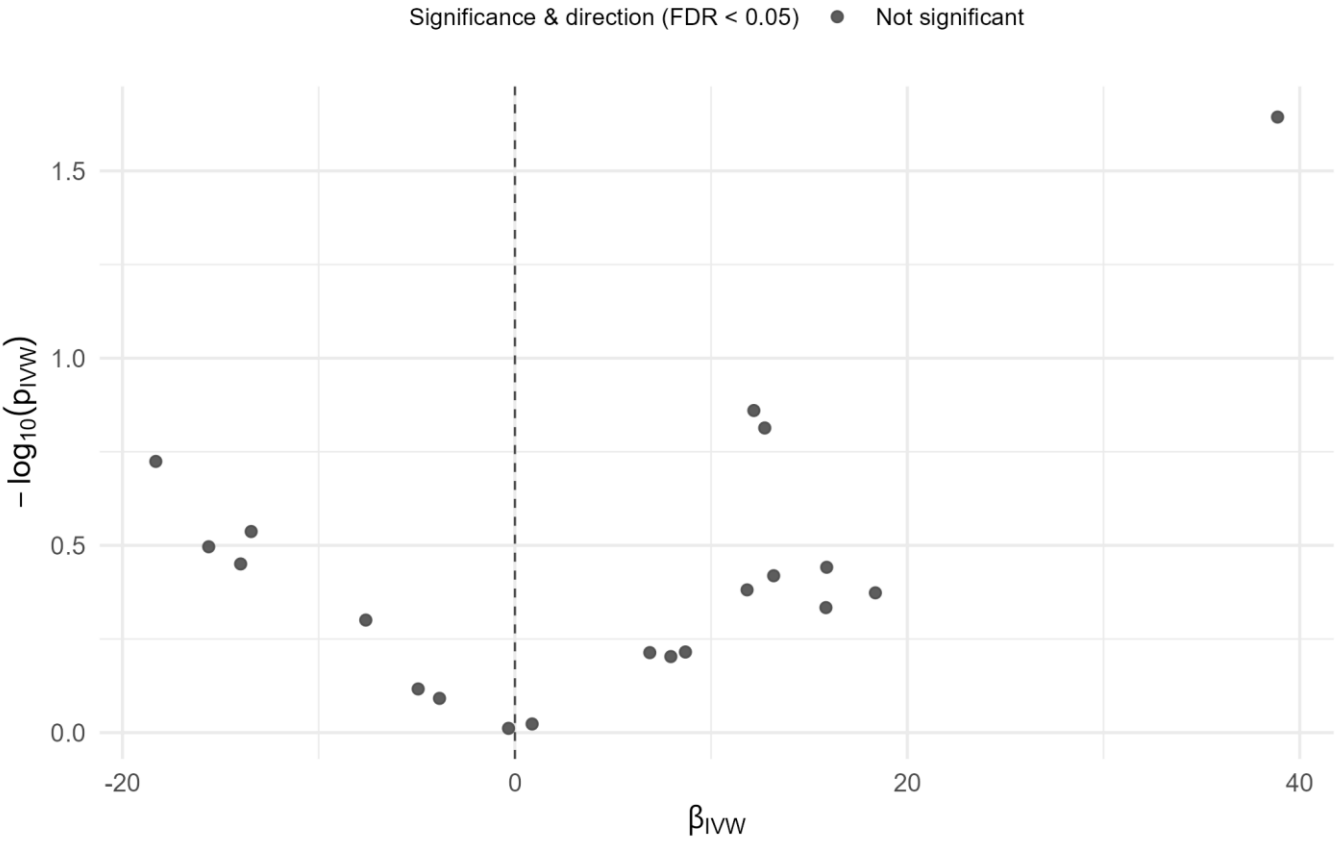


**Supplementary Figure 30.** Volcano plot results of phenome-wide Mendelian randomization of smoking initiation in both-sex East Asian population. "MR" refers to Mendelian Randomization testing. "smoking initiation" refers to genetically proxied smoking initiation. "FDR" refers to Benjamini-Hochberg false discovery rate correction. "βivw" refers to the causal effect of one standard deviation increase of smoking initiation on log-odds ratio of disease diagnosis as estimated using the inverse variance weighted method. The dotted line refers to a log-odds ratio of zero, implying no effect change in disease risk resulting from increased smoking initiation. Disease labels are applied to selected novel Mendelian randomization findings with selective labelling. Red dots imply increased smoking initiation raises risk for the named disease outcome, blue dots imply increased smoking initiation lowers risk, and grey dots represent null findings. Statistical significance is defined as $q < 0.05$ after Benjamini-Hochberg false discovery rate adjustment.
